# Quantifying Fine-scale Vegetation Associated with *Ixodes scapularis* Density to Inform Public Health Programming, Western Massachusetts

**DOI:** 10.64898/2026.10.14.26363243

**Authors:** James Tsalah, Veronica Bobskill, Johanna Ravenhurst, Kristen A. Ross, Allison M. Gardner, William J. Landesman, Andrew A. Lover

## Abstract

Vegetation plays a critical role in shaping fine-scale tick density by structuring local microclimates, habitat resources, and host interactions that largely determine tick survival and activity, with implications for tick-borne disease transmission. Tick burden in the United States continues to increase, with annual reported tick-borne disease cases more than doubling from 2004 through 2019. These trends call for a broader scope of tick-mitigation strategies that go beyond traditional prevention measures that focus on personal protection; and there is an opportunity to inform habitat-level interventions. Here, we quantify fine-scale vegetation structure across 48 subplots at tick surveillance sites in Western Massachusetts to assess local habitat features associated with *Ixodes scapularis* density and potential for human-tick contact risk. In assessing understory and overstory vegetation attributes, this study identified life stage-specific differences in predicted tick density, with higher adult densities in areas with deeper leaf litter and presence of invasive plant species and nymphs associated with high presence of ferns. Both adult and nymphal *Ix. scapularis* were predicted at higher densities in relatively low basal area forests (18.9 to 26.9 m²/hectare). This study design provides a simple, low-cost framework for organizations looking to incorporate tick-informed forest measurement ecology into their public health tick surveillance protocols with minimal training.

## 1. Introduction

The public health impact of tick-borne disease (TBD) has been increasing in recent decades across many temperate areas, including the United States (U.S.).^1^ In the northeastern U.S., the blacklegged tick (*Ixodes scapularis)* is the dominant vector of human disease agents and is closely tied to forested areas.^2^ The ecological drivers of tick survival and persistence operate across multiple spatial scales, and associations between tick population density and environmental/landscape factors have been demonstrated from the micro-environment (i.e. vegetation and leaf litter) to the macro-scale (forest-level). Survival is linked to quantifiable environmental factors, as *Ix. scapularis* spend the majority of their lives off-host, typically feeding on mammalian hosts for a maximum of 25 total days^3^ out of an approximate 2-year lifespan.^4^ However, there is limited consensus on what constitutes high-quality off-host habitat within acarological literature; furthermore direct comparisons are challenging due to variation in spatial scale.

Previous studies have examined a diverse set of factors influencing tick populations, including forest management, vegetation cover, overwintering survival conditions, forest fragmentation, elevation, and aspect.^5–12^ Micro-scale environmental factors such as leaf litter depth, shrubs, ferns, and understory vegetation cover have been associated with increasing *Ix*. *scapularis* density in temperate forests and parks.^8,10–13^ Macro-scale features including deciduous forest, closed forest canopies, forest fragmentation, and deciduous forest cover have also been associated with increasing *Ix. scapularis* density, while pines and grasslands have been associated with lower *Ix. scapularis* density.^5,13^

While personal protection is the mainstay for public health programming, a Centers for Disease Control and Prevention (CDC) survey found that over a third of respondents in New England do not engage in any personal protective measures,^14^ and a recent survey in the same region found that a majority of households self-reported only using personal protection “sometimes.”^15^ Consequently, there is a pressing need to incorporate other approaches, including landscape management strategies that may complement the “holes” in personal protection.^16^

While specific guidance exists for land use managers to mitigate the risk of increasing TBD on federal lands stemming from invasive plant species,^17^ several gaps remain. First, this guidance is focused on federal agencies with extensive capacity for field surveys; and second, it provides only limited information on mitigation strategies beyond habitat restoration, which may be unfeasible or cost-prohibitive in many areas. Many local public health offices and individual municipalities have insufficient resources to conduct intensive vegetation analysis or long-term tick collection. Developing cost-effective and efficient strategies to quickly characterize recreational/forested areas of potential higher tick risk for targeted interventions would benefit both public land managers and local public health officials.

Using two years of tick collection data and a range of habitat-level, micro-scale vegetation indices at each site, this study determines associations between rapidly collectable vegetation metrics and tick densities. In our study, we define tick habitat as the 50-100 m scale area comprising all off-host space and environmental conditions in which ticks engage in their natural behaviors such as questing, molting, and diapause/quiescence. Associations were quantified with three separate multivariable models to inform the distinct goals of routine public health surveillance and ecological studies. Results are presented as a combined adult-nymphal model to inform routine public health surveillance; and as two life stage-specific models to inform ecological studies.

## 2. Materials and Methods

### 2.1 Study area and site selection

The Pioneer Valley in Western Massachusetts, U.S., consists of the towns that surround the Connecticut River across three counties (Franklin, Hampden, Hampshire). The region is a complex landscape of paved development, suburban homes, second growth forest, and widespread agrarian sites. Agricultural and other extractive land practices prior to the early 20th century resulted in major forest loss,^18,19^ but as of 2017, over 50% of Massachusetts remains forested, the majority of which is privately owned.^20^ These southern New England forests are dominated by red maple (*Acer rubrum*), eastern white pine (*Pinus strobus*), northern red oak (*Quercus rubra*), and eastern hemlock (*Tsuga canadensis*).^20^ Our study sites reflect this composition, with most sited in oak-pine forests; specifically white pine/northern red oak/red maple (Table 1). Common mammalian hosts at these sites include the white-footed mouse (*Peromycus leucopus*), eastern chipmunk (*Tamais striatus*), northern short-tailed shrew (*Blarina brevicauda)*, woodland vole (*Microtus pinetorum*), and white-tailed deer (*Odocoileus virginianus*).^21–23^

**Table 1.** Overview of field sampling sites, Western Massachusetts, 2024-2025.

| Plot | Size | Description |
| --- | --- | --- |
| A | 40x40 | Eastern hemlock forest with an understory of American witch-hazel, mountain laurel and blueberry. High prevalence of coarse woody debris. A popular hiking trail bisects the tick collection plot. |
| B | 70x10 | Half of the plot is Oak - Hemlock - White pine forest, dominated by northern red oaks. Understory contains blueberry, eastern white pine, red maple, and glossy buckthorn. The other half of the plot is a powerline right-of-way, frequently cut with no overstory trees. Vegetation present includes hay-scented fern and dense herbaceous and woody vegetation. Near a hiking trail and adjacent to agricultural fields. |
| C1 | 40x40 | Chestnut oak forest, dominated by chestnut oaks and northern red oaks. Understory dominated by red maples, eastern white pine, and American witch-hazel. Scattered patches of lowbush blueberry and mountain laurel. Hiking trails pass through the plot. |
| C2 | 40x40 | Comparable to C1. Trails that intersect this plot are commonly used by mountain bikers |
| D1 | 40x40 | Oak - Hemlock - White pine forest, primarily dominated by northern red oak, with an understory of red maple and American witch-hazel with a dense covering of mountain laurel. |
| D2 | 60x20 | Hemlock forest with very little understory. Heavy soil compaction and many trails present. Adjacent to a popular swimming site and a paved bike trail. |
| E1 | 40x40 | Oak - Hemlock - White pine forest, overstory largely containing northern red oaks, eastern white pines. Ferns and lowbush blueberries dominate the forest floor. Near hiking trails. |
| E2 | 40x40 | Oak - Hemlock - White pine forest, containing eastern white pines, northern red oaks, and some birch with scattered patches of understory mountain laurel. High prevalence of coarse woody debris. Near hiking trails. |
| F | 40x40 | Successional northern hardwood forest, dominated by overstory red maple and black birch. Understory vegetation includes black cherry, shagback hickory, and red maple. Some ferns and lowbush blueberries are present. Located adjacent to a high-use accessible hiking trail. |

Communities throughout the Northeastern U.S. have extensive TBD burden: in 2023, tick-borne infections affected over 77,000 people in the Northeast US, with over 23% of those cases in southern New England (i.e., Connecticut, Massachusetts, Rhode Island).^24^ According to state-level reporting, western Massachusetts (Berkshire, Franklin, Hampden, and Hampshire counties) accounted for 8.4% of tick exposure emergency department visits during the same time period,^25^ with about 11.8% of the state population.^26^

To identify tick-borne disease hotspots in the region and select study sites, human movement data from an unpublished prospective spatial case control study^27^ in clinical patients was analyzed using quantitative cluster detection in SaTScan.^28^ GPS-derived movement data was collected from participants who had attached ticks before or during the study, and a control group with no self-reported recent history of attached ticks. Spatiotemporal clustering of case individuals was assessed with a retrospective space-time permutation model to detect statistically significant clusters of case occurrences.^29^ Analyses were conducted across multiple seasonal groupings and case-to-control ratios to provide “consensus” tick exposure hotspots.

These hotspots were evaluated for suitability as tick surveillance sites using GIS maps and site visits, which considered accessibility, potential for flooding, landowner permission for sampling, and feasibility for conducting tick dragging on 40×40 meter plots over multiple seasons. Six sites were selected in Hampshire County, MA (Figure 1A).

**Figure 1.**
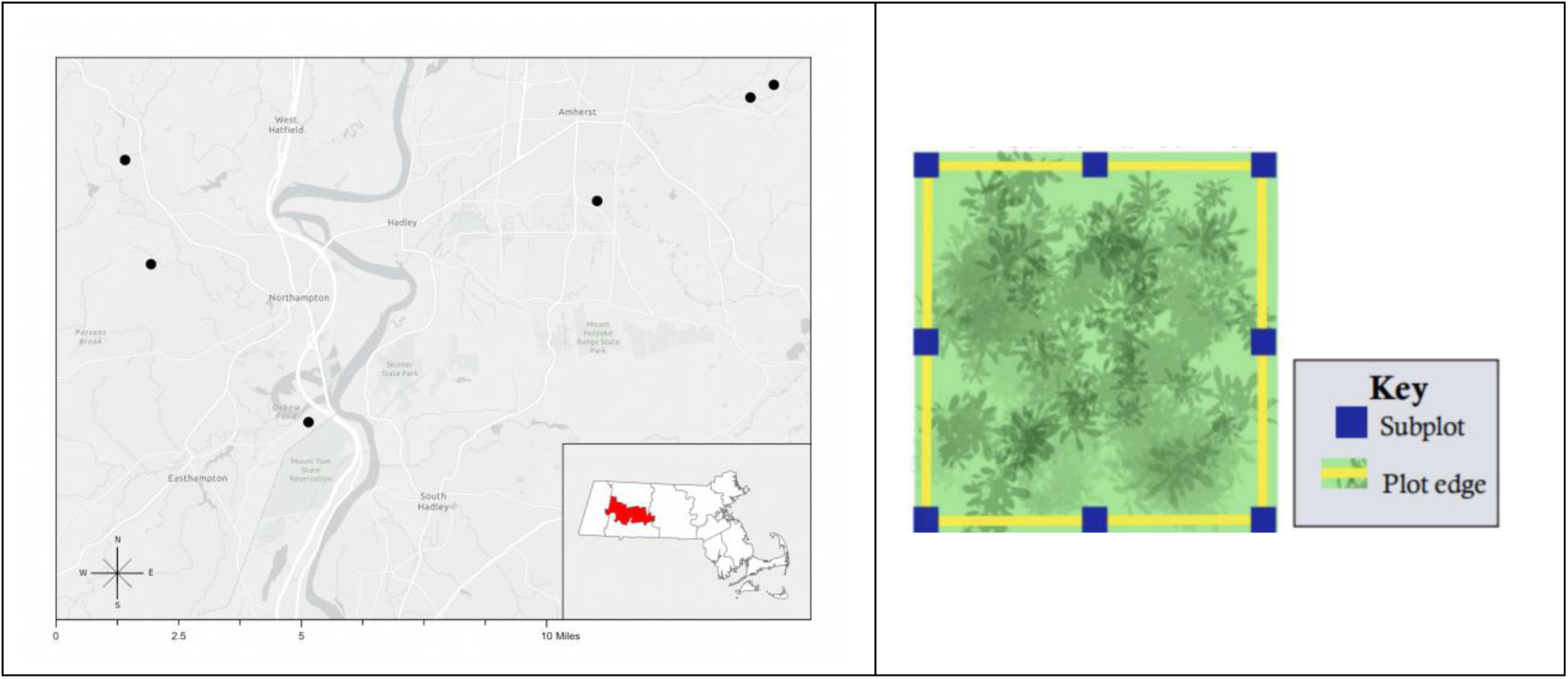
Panel. **A.** Location of field sampling sites in Hampshire County, Massachusetts. 2024-2025. Basemap reference is Light Grey Canvas Map (2026).^33^ **Panel B**. Schematic of tick and vegetation plot survey plan, Western Massachusetts, 2024-2025.

### 2.2 Plot Configurations & Sampling

After identification of suitable sites, study-plot grids were established. Vegetation surveys were performed at eight 3×3m subplots every 20m within overall 40×40m tick collection plots (Figure 1B). Vegetation surveys were conducted during August-October, prior to leaf fall, with a single sampling event per plot. Seven out of the nine vegetation surveys were conducted in 2024, with the remaining two late addition tick surveillance plots being surveyed in 2025. These surveys captured both understory and overstory indices, aligned with previous studies.^5,12^ Specifically, measurements of the overstory included basal area, percent canopy coverage, and presence of mast species (*Quercus* spp./*Juglans* spp./*Carya* spp.). Basal area is the total cross-sectional surface area per hectare if each live tree was cut at its diameter at breast height (DBH). To calculate it, we used a variable-radius plot method, which only requires setting a plot center, in contrast to fixed-radius plots, which would need a measured perimeter prior to finding basal area. To calculate basal area, the sampler stood at the center of the subplot, and pivoted 365° while peering through the window of an angle gauge using a basal area factor of 10 (Jim-Gem Cruz-All, PN 59795). Percent canopy coverage was assessed using a densitometer (GRS model# D0001) at eight unique, equidistant points along the perimeter of each subplot. Based on the total of positive canopy measurements, the percent canopy cover was calculated as per manufacturer’s instructions.

Leaf litter depth was measured twice per subplot using a soil knife, and categorized as deciduous, coniferous, or mixed. The height of the tallest plant was categorized as < 0.5m, 0.5-1.0m, or > 1.0m. Invasive plant species were described on a 0-3 scale, ranging from “none present” to “subplot entirely filled”. Specific invasive plant species were noted, including Asiatic bittersweet (*Celastrus orbiculatus*), barberry (*Berberis* spp.), black locust (*Robinia pseudoacacia*), Eurasian honeysuckle (*Lonicera* spp.) garlic mustard (*Alliaria petiolata*), glossy buckthorn (*Frangula alnus*), common buckthorn (*Rhamnus cathartica*), multiflora rose (*Rosa multiflora)*, Norway maple (*Acer platanoides*), purple loosestrife (*Lythrum salicaria*), burning-bush (*Euonymus alatus*), among others.

Understory presence/absence categories included berry species (*Vaccinium* spp./*Rubus* spp.), coarse woody debris, short and tall brushy vegetation, tall grasses, forbs, and ferns. Short brushy vegetation was a dense cover of short (< 76.2cm) shrubs that are easily passable and tall brushy vegetation was a dense cover of tall shrubs which obstructed free passage. The understory vegetation coverage was assessed on an ordinal 0-3 scale, including both native and non-native species. Additional details are in supplemental materials (Table S3; Supplemental files 1 and 2 - vegetation survey collection sheet & protocol).

Ambient site temperature and relative humidity were recorded with a data logger (HOBO MX2300) at each tick sampling event; units were mounted permanently within a solar radiation shield, logging data hourly. Cloud coverage and general weather conditions were recorded manually at each tick sampling event.

Tick collection in 2024 occurred approximately every two weeks from June 25 th through August 5th or nymphs, with no collections made during larval season, and resumed October 21 through November 25 for adults (2-7 total tick collections per site). Collections in 2025 took place from Mar 19 - Aug 4; and Sept 30 - Nov 24 (13 total tick collections per site). Site selection and placement in 2024 allowed for earlier sampling events in 2025. ( Table 3; Table S1) A total of nine 40×40m tick collection plots within six field sampling sites were used throughout the study period. For tick collections, a 1-m^2^ rubberized flannel drag cloth was used to sample the perimeter of the plot, stopping every 10m to inspect the cloth for nymph or adult stage ticks and discarding tick larvae.^30^ Sampling avoided dense thorny vegetation, wetlands, or poison ivy when possible. Collected ticks were stored in 70% ethanol, and stored at 4°C. Later, field-identified ticks were identified to species under a stereo microscope using standard taxonomic keys.^31,32^

### 2.3 Data processing

Vegetation metrics from subplots were aggregated to capture overall plot characteristics, using measures appropriate for each data distribution. Distributions were assessed with Shapiro–Wilk tests for normality and generated Q–Q plots to visually assess deviations from Gaussian distributions. Variables with W-statistics near unity and non-significant p-values (p ≥ 0.05) were classified as normal, while those deviating significantly from normality were categorized as skewed. To allow for interpretable estimates for continuous metrics, optimal data driven-breakpoints were determined using k-means (*discretize* within the *arules* package,^34^ testing 2-4 breakpoints and also as presence/absence), with final model choice guided via AIC. Metrics with approximately Gaussian distributions (Leaf Litter Depth, Understory Vegetation Density, Temperature, and Humidity) along with binary variables were summarized using means; skewed variables, using medians (Percent Canopy Cover, Invasive Species Density); and categorical variables using modal values (Basal Area, Leaf Litter Composition, Height of Tallest Plant/Shrub in Plot).

During model development, some characteristics were recategorized to improve interpretability of results, reduce collinearity, and optimize model AIC. Binary presence/absence variables included berry species, coarse woody debris, short and tall brushy vegetation, tall grasses, forbs, mast species, and invasive species. Invasive species (originally recorded on an ordinal 0 - 3 scale), was recategorized as a binary presence/absence to stabilize model performance.

Additionally, ordinal variables included understory vegetation density, height of tallest plant, ferns, and basal area. Plot-level fern measures were recategorized from presence/absence to “low density” (present in < 50% of subplots) and high density (present in ≥ 50% of subplots); while basal area, measured as a continuous variable (implying a continuous impact on tick density), was revised to an ordinal variable with three categories [35.2,43.1], [18.9,26.9], [26.9,35.2], with optimal breakpoints determined by k-means. (Table 2)

**Table 2.** Vegetation metrics recorded at sampling sites; Western Massachusetts, 2024-2025.

| Plot characteristic | Units | Scale; and Variable Transformations |
| --- | --- | --- |
| Leaf Litter Depth | cm | Continuous. |
| Basal Area | m<sup>2</sup>/hectare<br>18.9 - 26.9<br>26.9 - 35.2<br>35.2 - 43.1 | Continuous;<br>transformed to 3 categories via k-means |
| Ferns | Low (< 50%);<br>High (≥ 50%) | Presence/absence; transformed to low density<br>(present in < 50% of subplots) and high<br>density (present in ≥ 50% of subplots) |
| Temperature | °C | Continuous |
| Relative Humidity<br>(RH) | % | Continuous |
| Percent Canopy | % | Continuous |
| Invasive Species<br>Density | 0 to 3 | Ordinal; transformed to binary<br>presence/absence |
| Understory<br>Vegetation Density | 0 to 3 | Ordinal |
| Berry Species<br>(*Vaccinium*/*Rubus*) | 0/1 | Binary |
| Mast Species | 0/1 | Binary |
| Short Brushy<br>Vegetation | 0/1 | Binary |
| Tall Brushy<br>Vegetation | 0/1 | Binary |
| Height of Tallest Plant | None<br><0.5m<br>0.5 - 1m<br>>1m | Ordinal |
| Coarse Woody Debris | 0/1 | Binary |
| Tall Grasses | 0/1 | Binary |
| Forbs | 0/1 | Binary |

### 2.4 Multivariable models

Multilevel (“mixed”) negative binomial models were used to quantify ecological factors associated with tick density, using the R package *glmmTMB*.^35^ These models provide unbiased errors with the presence of repeated sampling and clustering within sites;^36^ site and sampling day were included as random effects.

Three independent *Ix. scapularis* models were constructed: nymphs-only; adults-only (to understand underlying ecological drivers) and nymph+adult combined (to support general surveillance activities). For each model, factors with p<0.20 in bivariate analysis were included in model building;^37^ Akaike Information Criteria (AIC) was used as the primary metric for model parsimony. To minimize collinearity between ecological metrics, model selection also used generalized variance inflation factor (VIF) from the R package *performance*.^38^ Many ecological measures showed extensive collinearity (Figure 2); in order to filter these unstable models, only models with a VIF value <3^39^ were examined further as separation^40^ was observed in this dataset at higher VIF values. Model specification and adequacy were assessed using the R package *performance*;^38^ and the *DHARMa* package was used to evaluate simulated model residuals.^41^ Effect sizes are reported as exponentiated coefficients (providing incidence rate ratios). All models included temperature and relative humidity to address residual confounding. Analyses were performed with R Software, 4.4.2; all tests were two-tailed, with ɑ = 0.05.

**Figure 2.**
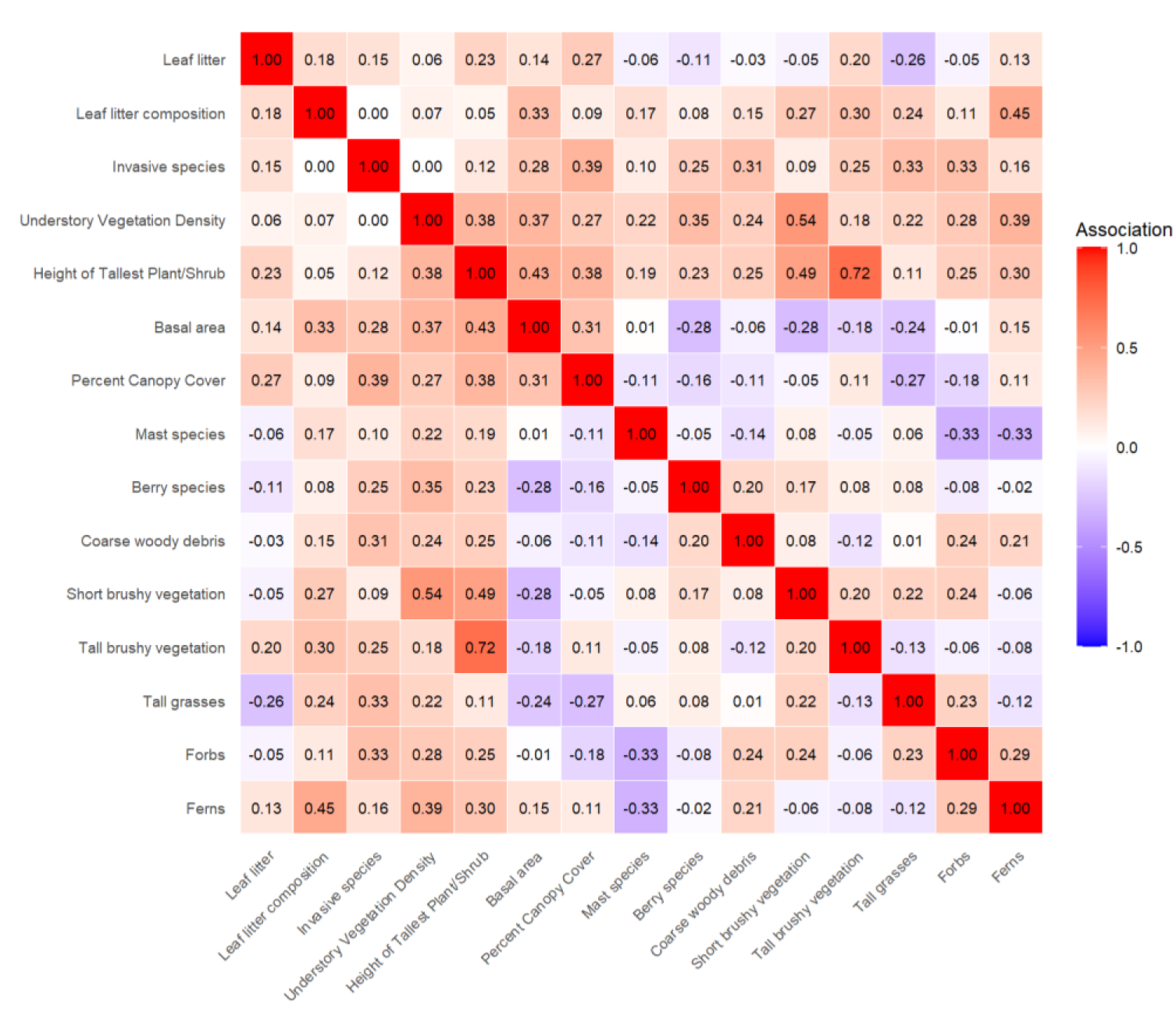
Mixed correlation plot of ecological indices, Western Massachusetts, 2024-2025. Note: Values normalized after calculation using different metrics depending on interaction: nominal vs nominal uses a bias-corrected Cramer’s V, nominal/numeric vs numeric with Spearman correlation.

## 3. Results

### 3.1 Descriptive analysis-tick densities

We conducted 155 quadrat-based tick drag sampling observations across six sites in Hampshire County, Massachusetts, between June 2024 and November 2025 ( Fig. 1A). A total of 622 *Ix. scapularis* ticks were collected with 241 nymphs and 371 adults (Table 3). Additionally, a total of 111 *Dermacentor variabilis* individuals were collected, all adults (Table 3).

**Table 3.** Overview values of the number of sampling events and ticks collected during Year 1 (2024) and Year 2 (2025) in Western Massachusetts. N/A values indicate that a site was not sampled in a given year.

| Plot | Sampling Events | Species | Stage | Total Year 1 | Total Year 2 | Total Ticks | Density per 100m<sup>2</sup> |
| --- | --- | --- | --- | --- | --- | --- | --- |
| A | 15 | *Ix. scapularis* | Adult | 2 | 15 | 55 | 2.3 |
|  |  |  | Nymph | 6 | 38 |  |  |
|  |  | *D. variabilis* | Adult | 0 | 0 |  |  |
| B | 18 | *Ix. scapularis* | Adult | 60 | 182 | 412 | 14.3 |
|  |  |  | Nymph | 6 | 63 |  |  |
|  |  | *D. variabilis* | Adult | 0 | 101 |  |  |
| C1 | 19 | *Ix. scapularis* | Adult | 1 | 33 | 59 | 1.9 |
|  |  |  | Nymph | 2 | 22 |  |  |
|  |  | *D. variabilis* | Adult | 0 | 1 |  |  |
| C2 | 19 | *Ix. scapularis* | Adult | 1 | 5 | 27 | 0.9 |
|  |  |  | Nymph | 2 | 19 |  |  |
|  |  | *D. variabilis* | Adult | 0 | 0 |  |  |
| D1 | 19 | *Ix. scapularis* | Adult | 4 | 16 | 26 | 0.8 |
|  |  |  | Nymph | 1 | 2 |  |  |
|  |  | *D. variabilis* | Adult | 0 | 3 |  |  |
| D2 | 19 | *Ix. scapularis* | Adult | 0 | 1 | 3 | 0.1 |
|  |  |  | Nymph | 0 | 2 |  |  |
|  |  | *D. variabilis* | Adult | 0 | 0 |  |  |
| E1 | 13 | *Ix. scapularis* | Adult | N/A | 25 | 77 | 3.7 |
|  |  |  | Nymph | N/A | 51 |  |  |
|  |  | *D. variabilis* | Adult | N/A | 1 |  |  |
| E2 | 13 | *Ix. scapularis* | Adult | N/A | 12 | 29 | 1.4 |
|  |  |  | Nymph | N/A | 16 |  |  |
|  |  | *D. variabilis* | Adult | N/A | 1 |  |  |
| F | 20 | *Ix. scapularis* | Adult | 7 | 7 | 30 | 0.9 |
|  |  |  | Nymph | 2 | 9 |  |  |
|  |  | *D. variabilis* | Adult | 2 | 3 |  |  |
| Totals | 155 | *Ix. scapularis* | Adult | 75 | 296 | 723 | N/A |
|  |  |  | Nymph | 19 | 222 |  |  |
|  |  | *D. variabilis* | Adult | 2 | 109 |  |  |

Tick density was highly heterogeneous across sampling locations: Site B accounted for the majority of collections, contributing 57.4% (412) of total *Ix. scapularis*, and also had the highest single-visit yield (56 specimens). At the other extreme, site D2 had the lowest density, with only three *Ix. scapularis* sampled across the entire study period (Table 3 & Figure S1). Tick species composition also varied among sites-notably, *D. variabilis* was entirely absent from two sites (C2 and D2), despite each site being surveyed on 19 occasions (Table 3).

### 3.2 Descriptive analysis-habitat metrics

The study sites varied in dominant understory vegetation; several sites (Plots A, E1, E2, F) were dominated by ferns while others (Plots A, B, C1, C2, D1) had complete coverage of short brushy vegetation. Sites also had several similarities-all shared the presence of mast species, short and tall brushy vegetation and coarse woody debris, albeit at different levels. Additionally, this study included several outlier sites with unique vegetation not reflected at other sites.

Specifically, Plot C2 had very high basal area relative to the others, with 43 m^2^/hectare, while Plot D2 contained less understory vegetation overall, including coarse woody debris, forbs, short and tall brushy vegetation. All sites had full canopy coverage except Site B (62.5%), which included a powerline easement within its sampling area, and also lies directly adjacent to wetlands and cornfields. Additional details on vegetation metrics by plot are in supplemental materials (Table S2 & Table S4).

In addition to site-specific trends, many of the measured ecological indices were strongly correlated with one another, showing both positive and negative associations. Figure 2 shows all pairwise comparisons of factors captured in the vegetation indices. This figure utilizes a range of comparisons between the data types, which have been normalized for comparisons. Several of the measures show few strong correlations (e.g. mast species; leaf litter), indicating these capture “unique” aspects of the ecology; while others (e.g. understory vegetation density; tall grasses; basal area) are highly correlated with many other measures. This indicates these metrics are all capturing subtly different aspects of the specific ecology at the study sites.

Specifically, basal area and percent canopy coverage had a positive association with understory-related metrics. The height of the tallest plant/shrub in the subplot was positively associated with basal area, and with tall and short brushy vegetation in subplots. Short brushy vegetation was positively associated with understory vegetation density, but had a negative association with basal area. Additionally, fern abundance was positively associated with understory vegetation density and leaf litter composition, while conversely ferns and forbs were negatively associated with mast species. These relationships are consistent with expected ecological trends-plant height and understory vegetation density are dependent on the presence of short and tall brushy vegetation, which is moderated by basal area and percent canopy cover.

### 3.3 Multivariable analyses

Three independent *Ix. scapularis* models were constructed: nymphs-only; adults-only (for ecological factors) and nymph+adult combined (for surveillance purposes). The distributions of variables included in the main models are shown in Figure 3; distribution of all other measured factors are shown in Figures S2-S4.

**Figure 3.**
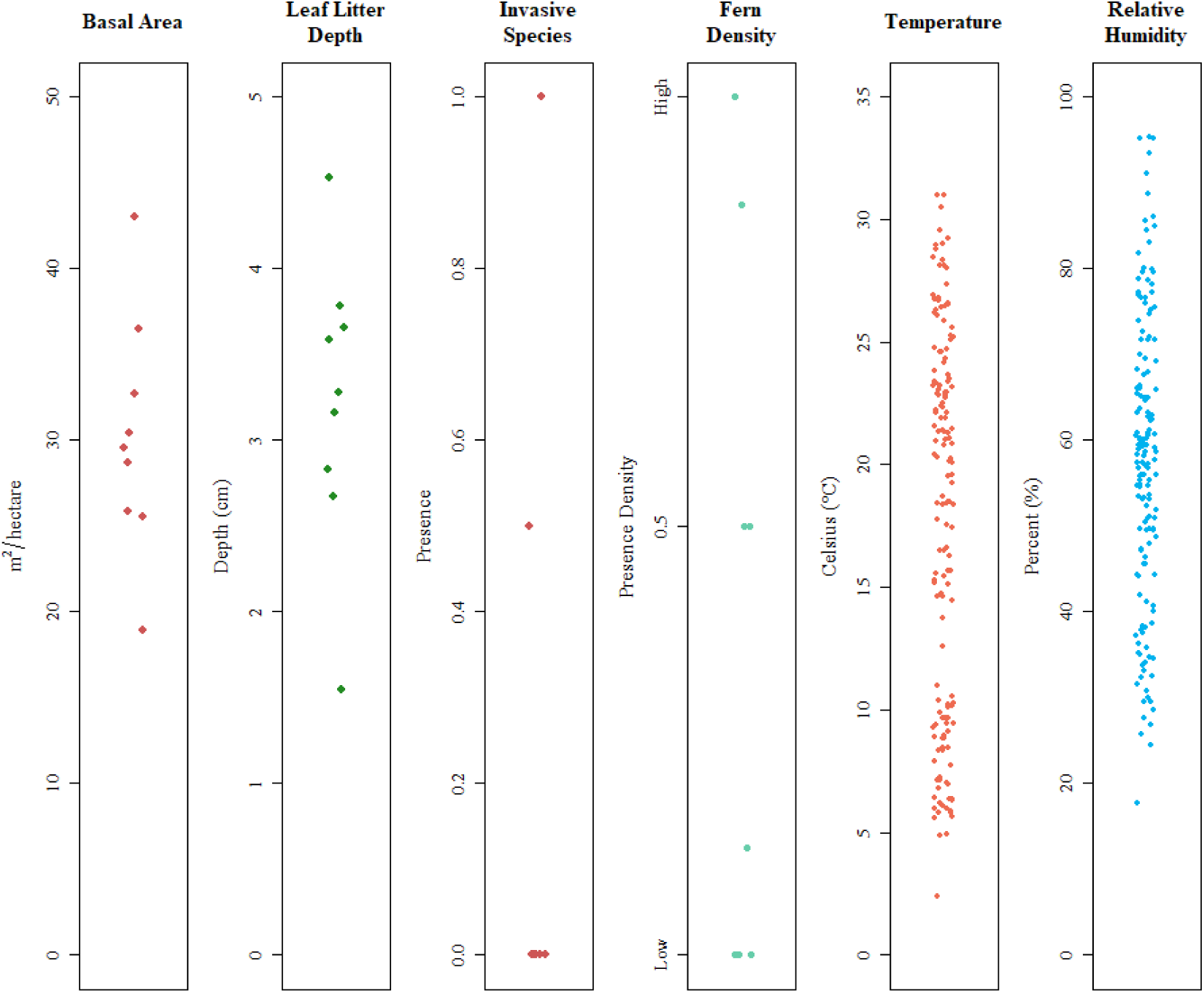
Distribution of plot-level variables included in final models, Western Massachusetts, 2024-2025.

#### Model 1. (Ix. scapularis, adults and nymphs combined)

In models including both adult and nymphal life stages (Table 4) leaf litter depth, fern density, and basal area had a statistically significant association with tick density after adjustment for temperature, relative humidity (RH), and year of sampling. Leaf litter depth and fern presence had positive associations with tick density: with leaf litter depth having an incidence rate ratio (IRR) of 1.64 (95% CI: 1.18 to 2.29; p < 0.001) while high fern density relative to low fern density had an IRR of 5.77 (95% CI: 3.70 to 8.98; p < 0.001). Conversely, there was a negative association between basal area and sampled *Ix. scapularis*. With the high basal area category (35.2,43.1 m²/ha) as reference, there was a positive association with sampled tick density with the intermediate basal area category (> 26.9 to 35.2 m²/ha) having an IRR of 2.15 (95% CI: 1.09 to 4.25, p = 0.027), while the low basal area category (18.9 to 26.9) had an IRR of 11.3 (95% CI: 5.61 to 22.7, p < 0.001).

**Table 4.** Mixed model negative binomial regression for association between landscape metrics and tick density (*Ix scapularis*, combined adults and nymphs), Western Massachusetts, 2024-2025. Notes: CI = Confidence Interval, IRR= incidence rate ratio.

| Characteristic | Category | IRR | 95% CI | p-value |
| --- | --- | --- | --- | --- |
| Leaf Litter Depth |  | 1.64 | 1.18, 2.29 | <0.001 |
| Basal Area (m<sup>2</sup>/ha) | High ( > 35.2 to 43.1) | reference | - | - |
|  | Medium ( > 26.9 to 35.2) | 2.15 | 1.09, 4.25 | 0.027 |
|  | Low (18.9 to 26.9) | 11.30 | 5.61, 22.7 | < 0.001 |
| Fern density | Low (< 50%) | reference | - | - |
| | High (≥ 50%) | 5.77 | 3.70, 8.98 | < 0.001 |
| Year | 2024 | reference | - | - |
|  | 2025 | 1.98 | 1.17, 8.98 | 0.011 |
| Relative humidity (%) |  | 1.01 | 1.00, 1.03 | 0.045 |
| Temperature (°C) |  | 1.00 | 0.97, 1.02 | 0.9 |

These results indicate that each centimeter of increasing leaf litter is independently associated with a 1.6-fold increase in sampled tick density, and high-density fern areas have 5.8-higher density of *Ix. scapularis* relative to low-fern sites. Additionally, categories of decreasing tree density are associated with 2.2-fold and 11.3-fold, higher tick numbers relative to the highest basal area sites within the study area. Lastly, there was an independent association with 2025 having higher tick density relative to 2024. No other measured factors had a statistically significant association with tick density in this model.

#### Model 2. (Ix. scapularis nymphs-only)

In models for nymphal *lx. scapularis* density (Table 5), fern density and basal area were found to have statistically significant associations with tick density, but with reduced effect sizes compared to *Model 1*. Results were adjusted for temperature (IRR = 1.16; 95%CI: 1.10 to 1.22; p< 0.001), relative humidity (not significant), and year. High fern density relative to low fern density (3.60; 95% CI: 1.82 to 7.10; p = < 0.001) had positive associations with tick density, while there was a negative association between basal area and sampled nymphs. With the high basal area category as reference, there was no statistically significant association with nymphal density at the intermediate basal area category with an IRR of 1.11 (95% CI: 0.44 to 2.78; p = 0.8); but a significant association with the low basal area category areas with an IRR of 4.14 (95% CI: 1.56 to 11.0; p = 0.004).

**Table 5.** Mixed model negative binomial regression for association between landscape metrics and tick density (*Ix. scapularis*, nymphs only), Western Massachusetts 2024-2025. Notes: CI = Confidence Interval, IRR= incidence rate ratio.

| Characteristic | Category | IRR | 95% CI | p-value |
| --- | --- | --- | --- | --- |
| Basal Area (m<sup>2</sup>/ha) | High (> 35.2 to 43.1) | reference | - | - |
|  | Medium (> 26.9 to 35.2) | 1.11 | 0.44, 2.78 | 0.8 |
|  | Low (18.9 to 26.9) | 4.14 | 1.56, 11.0 | 0.004 |
| Fern density | Low (< 50%) | reference | - | - |
| | High ($\geq 50\%$) | 3.60 | 1.82, 7.10 | < 0.001 |
| Year | 2024 | reference | - | - |
|  | 2025 | 3.73 | 1.71, 8.12 | < 0.001 |
| Relative humidity (%) |  | 1.02 | 1.00, 1.05 | 0.48 |
| Temperature (°C) |  | 1.16 | 1.10, 1.22 | <0.001 |

The results of this model indicate that high density fern areas were associated with a 3.6 -higher density of nymphs relative to low-fern sites; and that low basal area category of tree density is associated with 4.14 -fold higher sampled nymphal numbers relative to the highest tree density sites within the study area. Year of sampling showed a larger impact relative to the adult-only model and 2025 had a 3.7-fold higher incidence rate relative to 2024, adjusted for all other factors in the model.

#### Model 3. (Ix. scapularis adults)

Models for adult *lx. scapularis* (Table 6) identified contrasting factors associated with tick density in comparison to models that included nymphs. Adult density retained the positive association with leaf litter depth (IRR = 4.19; 95% CI: 2.52 to 6.96; p < 0.001) seen in *Model 1*, and had a positive association with the presence of invasive species (11.8; 95% CI: 5.01 to 27.7; p < 0.001). With the high basal area category as reference, there was a statistically significant association with tick density: the intermediate basal area category has an IRR of 4.25 (95% CI: 1.48 to 12.2, p = 0.007), while the low basal area category had an IRR of 6.47 (95% CI: 2.24 to 18.7, p < 0.001).

**Table 6.**
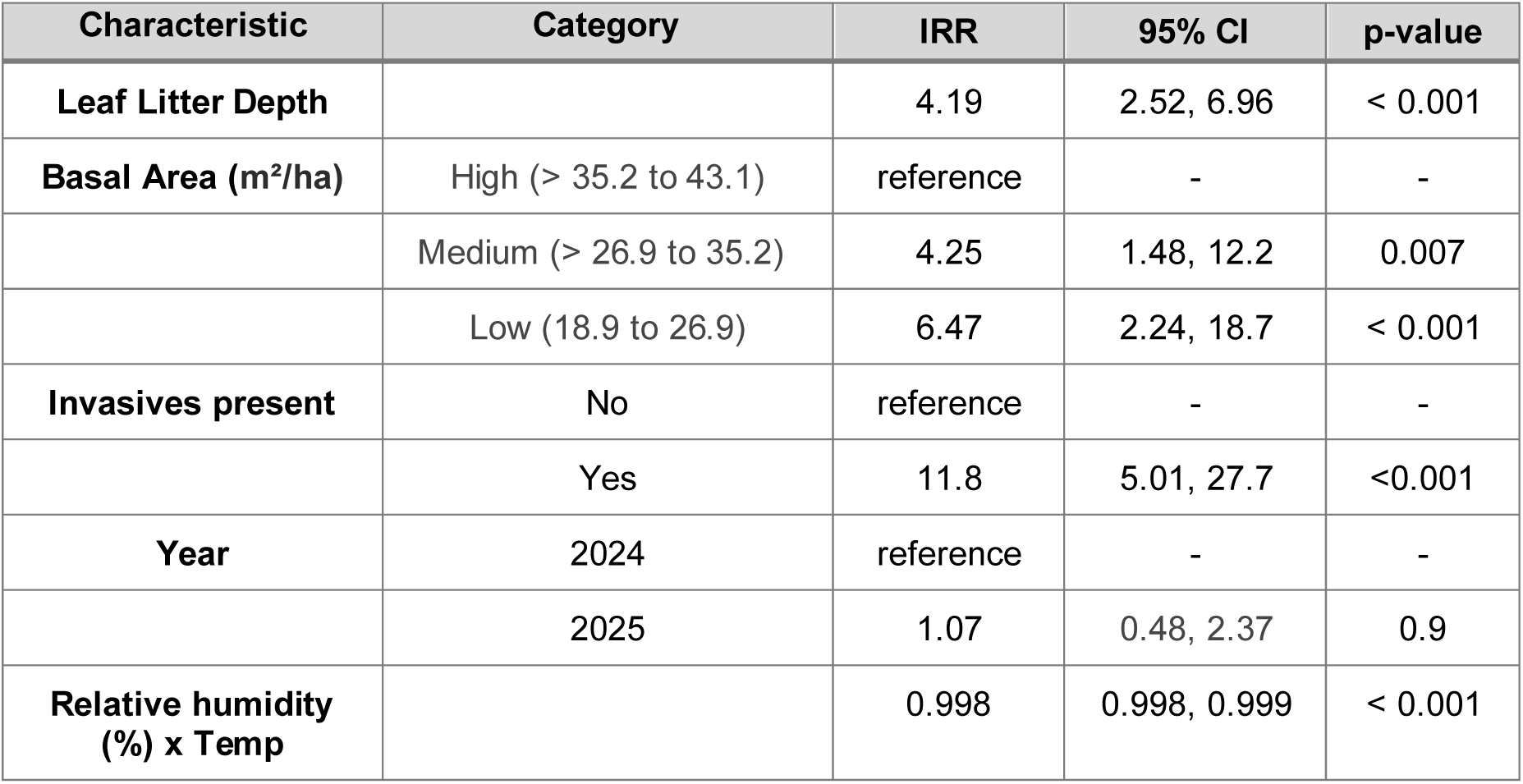
Mixed model negative binomial regression for association between landscape metrics and tick density (*Ix. scapularis*, adults only), Western Massachusetts, 2024-2025. (Notes: CI = Confidence Interval, IRR= incidence rate ratio).

| Characteristic | Category | IRR | 95% CI | p-value |
| --- | --- | --- | --- | --- |
| Leaf Litter Depth |  | 4.19 | 2.52, 6.96 | < 0.001 |
| Basal Area (m<sup>2</sup>/ha) | High (> 35.2 to 43.1) | reference | - | - |
|  | Medium (> 26.9 to 35.2) | 4.25 | 1.48, 12.2 | 0.007 |
|  | Low (18.9 to 26.9) | 6.47 | 2.24, 18.7 | < 0.001 |
| Invasives present | No | reference | - | - |
|  | Yes | 11.8 | 5.01, 27.7 | <0.001 |
| Year | 2024 | reference | - | - |
|  | 2025 | 1.07 | 0.48, 2.37 | 0.9 |
| Relative humidity (%) x Temp |  | 0.998 | 0.998, 0.999 | < 0.001 |

These results indicate that each 1-cm increase in leaf litter depth is associated with a 4.2-fold increase in adults; and the presence of invasive species is independently associated with an 11.8-fold increase in adult density. The middle basal area category was associated with a 4.3-fold greater rate of tick collection, while the lowest density areas had a 6.5-fold higher adult density relative to the densest forests in the study area. There was no statistically significant difference in the sampling rate comparing 2025 with 2024 in adjusted models.

## 4. Discussion

This study’s results quantify the impact of fine-scale variation across forest vegetation structures on tick abundance, highlighting opportunities to directly impact microhabitat conditions for vector control. Vegetation predictions varied between the life-stage specific models and the combined life-stage models. For the ‘‘nymphal stage” model, leaf litter depth was not associated with density, while fern density had a positive association. Conversely, in the “adult only” model, leaf litter depth and invasive species presence was positively associated with density while fern density did not show a statistically significant relationship. This contrast in ecological niche may be due to physiological differences between lifestages; adult *Ix. scapularis* are much more resistant to desiccation than nymphs.^42^ Across all models, predicted *lx. scapularis* density declined with increasing basal area. Although some of the aforementioned vegetation indices were consistent across models, the magnitude of effect varied between life stages.^43^ Increased leaf litter has been associated with higher tick densities in previous studies,^9–12^ and our findings are consistent with these observed patterns in adult ticks. However, leaf litter depth did not have a significant association with nymphal density in our study plots. Our findings are consistent with studies in New Jersey which suggest complex dynamics along shrub-leaf litter interfaces, where nymphs migrate to find optimal moisture regimes, while adults are less prone to move along these gradients.^44^ The specific ecology of our study sites (all had at least minimal leaf litter) could potentially mask any associations.

Fern presence may contribute to suitable tick habitat by increasing ground-level shading and maintaining local moisture conditions. Related studies in Maine (with a different analytical framework) reported positive associations with ferns.^11^ In this study, the positive association with high fern density was seen in the combined life stages and nymph models (IRRs of 5.77 and 3.60 respectively), but not with the adult model alone. This suggests that nymphs may benefit from the combined microclimate and questing structure effects of ferns within the understory during the warmer summer months, but these structures may be less impactful for adult ticks during the cooler fall sampling season. However, our data are unable to separate these two processes. Seasonal climatic shifts may also have a role in habitat preference and activity; the nymph population peaks in late spring and early summer, while adults are most active in the cooler fall and early spring months.^45^ Alternatively, these patterns may reflect life stage-specific differences in capture probability within fern-dominated habitats, driven by changes in behavior and questing capacity of *Ix. scapularis*.

In contrast with previous studies^12,46,47^ which did not find a significant association, or found a positive correlation between tick density and basal area, our study found lower basal area was associated with higher predicted *Ix. scapularis* density within mixed deciduous-coniferous forests. This may reflect indirect effects of stand structure on understory development, as higher basal area has been associated with reduced understory vegetation,^12^ potentially limiting host– tick encounter rates. In this study, basal area produced highly consistent effect estimates across models and the observed effect gradient of basal area is striking. This “dose dependence” of tick density with specific categories of timber density strengthens the proposed biological rationale for the observed decades-long increases in tick density, and suggests that there is an “optimal” tree stand density that supports *Ix. scapularis* ecology. This is consistent with studies in *Ixodes persulcatus* (the Eurasian taiga tick) showing maximum densities 12-25 years after forest clearing, with a gradual decay out to > 80 years post-logging.^48^

In comparable studies exploring the associations of basal area on tick densities, study design variations make direct comparisons difficult. Many of these studies focused on disturbed/logged sites and impacts of ecological succession on tick density, whereas our study sites are less dynamic.^7,47,49^ In addition, other studies included only the nymph stage,^7,12^ which could influence the results, as we found higher IRR values in both the adult-only and combined life stage models compared to nymph-only models. Additionally, previous studies used mean tick densities as a response variable^12^ compared with the unaggregated tick counts used in this analysis. These details suggest important nuances in assessing the impact of basal area in different settings.

Invasive species density was retained only in the adult-stage model, suggesting a life stage-specific relationship with adult tick abundance. The absence of this relationship in the nymphal life stage model suggests that invasive vegetation may be linked more strongly to the ecological processes affecting adult ticks than to factors influencing immature stages. Since invasive vegetation can influence both microclimate and host species composition, disentangling direct vegetation effects from broader habitat processes will require more detailed studies over a longer time period. Prior studies have identified several species of note in this regard, especially Japanese barberry^8,11,50^ and Eurasian honeysuckle.^8,11^

Despite extensive consideration in the ecology literature, variation in spatial scales makes comparison between studies challenging. Environmental data regarding ticks has been assessed on scales from 1m^2^ for microclimate and understory studies, all the way to >5km^2^ for landscape composition.^49^ Different forest types,^51^ soil classifications, climates and host dynamics^52^ may also further obfuscate trends. Recent work modeling the effect of spatial scales on density of *Ix. scapularis* nymphs (DON) showed greatest exploratory power at smallest scales; DON was most accurately predicted at a scale of 100m.^53^ However, with varying methods of data collection and spatial aggregation, it is unclear what scale may be best for predicting tick density and overall TBD transmission. Although these results are promising, more research would have to be conducted to determine what level of observation and variables are most impactful. Examining determinants of “pathogenic landscapes”, small-scale tick habitat features, and human land use patterns in the context of tick density patterns will help us create further interventions to complement personal protection.^54^ While this is a key goal of practical public health, our study design cannot assess the potential causal impact of vegetation management.

The inclusion of more factors may be one reason for why this work identifies different associations than other studies. By considering a broad range of vegetation characteristics, some variables may describe similar effects on tick density and were therefore removed from consideration to avoid including factors that were too closely related. Expanding habitat scope thus requires more statistically rigorous ecological data analysis to more holistically evaluate the complex relationships between ticks and their environments. Prior studies assessing relationships between vegetation characteristics and tick density have historically relied on standard GLM,^55^ GLMM,^56^ or zero-inflated^57^ multivariable models. As ecological modelling approaches continue to develop, adoption of modern analytical methods can support emerging study designs. Our analysis evaluates many vegetation metrics to characterize ecosystem-scale effects that may be difficult to capture using some previous approaches. Distinct from similar methods used in Conte et al.^7^ which used generalized linear mixed models implemented in the *glmmTMB* modeling package,^35^ inferences in our study are made at the scale of aggregated vegetation conditions rather than individual observations, and the reported incident rate ratios reflect this level of analysis.

The vegetation metrics developed in this study were designed to be simple and feasible in most settings, and require minimal training without sacrificing analytical utility. A 2024 survey of land trust organizations across the northeast and midwestern United States found that 36% of respondents used vegetation modification techniques, but a lack of trained personnel accounted for a significant barrier to increasing tick surveillance or control activities.^58^ This study used a simple 0-3 ordinal scale and binary presence/absence for most vegetation metrics to reduce invested time while still recording key vegetation features and sufficient replicants. In mixed deciduous-coniferous forests in New England, our evidence supports that basal area seems to be a strong predictor of risk. Basal area is a metric that can easily be incorporated into routine entomological surveillance-a simple metal angle gauge ($20 USD) can be used with minimal training. This can provide a straightforward way for health districts and land managers to assess potential tick exposure risks. The associations found between leaf litter, fern cover, and tick density suggest that these habitat characteristics may provide practical indicators for landscape-scale surveillance. These variables can be quantified quickly in the field and require minimal equipment to facilitate a standardized assessment across potential sites toward sustainable tick control. However, before basal area can be considered as a summary metric for acarological risk, it should be assessed in a wider range of ecologies and across different forest succession trajectories.

While our study provides important insight into the association between vegetation and tick density, there are several limitations that may impact our conclusions. Firstly, fixed quadrilateral plots in our study made placement within the greater landscape challenging. Although we had many potential sampling locations from our previous spatial hotspot analysis, many were ultimately excluded due to extensive wetlands, poison ivy or thorny plants that overlapped with our large square plot. These limitations mean other areas of highest risk may not have been included on the plots. Secondly, peak nymphal activity was not captured in 2024, but subsequently was in 2025. Thirdly, tick sampling using dragging/flagging techniques may be influenced by local climate, environmental conditions, and dragging speed: with slower speeds yielding higher nymph collections compared to faster sampling speeds.^59^ Standardized sampling protocols for this work minimize these differences. However, these caveats compound as *single-drag per event* sampling, as used in this study, was estimated to capture approximately only 5.9 to 6.3% of the nymphal population.^59^ Finally, vegetation indices were generally characterized using a binary Presence/Absence framework for efficient field activities. As a result, this allowed for sampling for multiple subplots per tick collection plot, providing more comprehensive measures of the micro-ecology. Our binary collection provides a directly interpretable estimate for associations; however, it may obscure ecologically important relationships where thresholds may exist.

The factors identified here as being most strongly associated with tick density (basal area; presence of ferns; and invasive species) suggest multiple avenues for public health programs, including vegetation management in high usage areas; signage within higher-risk ecologies; and public British Broadcasting Corporation campaigns highlighting areas and times of highest risk. To date, studies have shown large impacts on nymphal density after removal of invasives over a 5-year sample period in the Midwestern US,^60^ but trials in other ecologies and assessing epidemiological outcomes should be a key priority. There have also been few studies evaluating the impact of “risk area” signage on reducing tick exposures, and findings from these are mixed.^61^ Operational research to optimize risk messaging, including trailhead signage for public health outcomes in a range of settings is an urgent need, as studies suggest different demographic groups may require specific, targeted communications for behavior change.^62,63^

While our study site locations were determined by human usage patterns, this work was unable to include quantifiable metrics for land use. Integrating human behavior and land usage patterns with forest cover metrics is a critical area to understand epidemiologically highest-risk areas.^64^ In contrast to other vector-borne diseases like malaria, where sustained control and prevention strategies have been central to mitigation of transmission, tick-borne disease management has historically prioritized research and personal protection over large-scale interventions. Landscape-scale focused vegetation management strategies may represent an underutilized approach for reducing tick populations and associated human infections.^65^

## Supporting information

Vegetation Survey Collection Sheet

Vegetation Survey Protocol

## Data Availability

All data produced in the present study are available upon reasonable request to the authors.

## Author Contributions statement

JT and VB are joint first authors.

AAL, AMG, WL, and KR contributed to the design of the vegetation survey. AAL, VB, JT, and JR contributed to the design and implementation of field protocols. VB led all field data collection.

JT and AAL analyzed the data. JT, VB, and AAL created the tables/figures and wrote the first draft. All authors contributed to revisions and finalization of the manuscript; AAL is the final guarantor for this work.

## Attributions

Maps were created using ArcGIS® software by Esri. ArcGIS® and ArcGIS Pro™ are the intellectual property of Esri and are used herein under license. Copyright © Esri. All rights reserved. For more information about Esri® software, please visit www.esri.com.

## Funding statement

Funding has been provided to NEWVEC/UMass Amherst by the US Centers for Disease Control and Prevention (cooperative agreement U01CK000661). Human data collection for the spatial case control was funded by NIH/NIAID (R21AI149176-01A1 to AAL).

## Supplemental Information

**Table S1.** Overview of plot sampling dates.

| Plot | Year | Month |
| --- | --- | --- |
| A | 2024 | Nov - Nov |
|  | 2025 | Mar - Nov |
| B | 2024 | Jul - Nov |
|  | 2025 | Mar - Nov |
| C1 | 2024 | Jul - Nov |
|  | 2025 | Mar - Nov |
| C2 | 2024 | Jul - Nov |
|  | 2025 | Mar - Nov |
| D1 | 2024 | Jul - Nov |
|  | 2025 | Mar - Nov |
| D2 | 2024 | Jul - Nov |
|  | 2025 | Mar - Nov |
| E1 | 2024 | Apr - Nov |
|  | 2025 | Apr - Nov |
| E2 | 2024 | Apr - Nov |
|  | 2025 | Apr - Nov |
| F | 2024 | Jun - Nov |
|  | 2025 | Mar - Nov |

**Table S2.** Overview of vegetation indices by study plot (summary values from subplots) Western Massachusetts, 2024-2025.

| Site | Leaf Litter Depth (cm) | Basal Area (m<sup>2</sup>/ha) | Fern Coverage (%) | Invasives Presence | Temperature °C Mean + (Range) | Relative Humidity Mean + (Range) |
| --- | --- | --- | --- | --- | --- | --- |
| A | 4.5 | 36.4 | 0.9 | 0 | 16.3°<br>(4.9° - 29.2°) | 60.2%<br>(33.8% - 86.0%) |
| B | 3.3 | 18.9 | 0.1 | 1 | 17.8°<br>(5.8° - 30.5°) | 50.9%<br>(24.4% - 76.8%) |
| C1 | 3.1 | 30.4 | 0 | 0 | 18.33°<br>(7.1° - 29°) | 54.8%<br>(31.5% - 79.9%) |
| C2 | 2.7 | 43 | 0 | 0 | 18.6°<br>(6.8° - 28.8°) | 54.1%<br>(32.3% - 77.2%) |
| D1 | 3.6 | 25.5 | 0 | 0 | 16.5°<br>(5.9° - 28.1°) | 65.8%<br>(34.6% - 95.1%) |
| D2 | 2.8 | 25.8 | 0 | 0 | 16.02°<br>(4.9° - 28.1°) | 66.8%<br>(35% - 95.3%) |
| E1 | 3.8 | 28.7 | 0.5 | 0 | 17.3°<br>(5.8° - 31°) | 52%<br>(25.7% - 69.2%) |
| E2 | 3.6 | 32.7 | 1 | 0 | 17.6°<br>(5.6° - 31°) | 55.9%<br>(29.6% - 75.1%) |
| F | 1.5 | 29.5 | 0.5 | 1 | 20.2°<br>(2.4° - 27.3°) | 54.7 %<br>(17.6 - 84.8) |

**Table S3.** Presence/Absence Key. Details of vegetation indices, Western Massachusetts, 2024-2025.

| Characteristic | Details |
| --- | --- |
| Mast Species | Oak, hickory or walnut species of any age within the subplot (either leaning in and providing canopy coverage, or growing within the subplot) |
| Berry species | *Vaccinium* and *Rubus* species present within or leaning into the subplot |
| Coarse woody debris | Downed trees or large branches (at least 10cm in diameter and 1m in length) |
| Short brushy vegetation | Dense cover of short (< 0.76m) shrubs that are easily passable |
| Tall brushy vegetation | Dense cover of tall shrubs ($\geq 0.76$m) which obstructs free passage. May require flagging |
| Tall grasses | Grasses, rushes, sedges (graminoids) and other weeds measuring 8 inches or greater in height |
| Ground cover vegetation | Low-growing vegetation < 0.91m; fully covers ground in 50% or more of area being evaluated |
| Forbs | Nongrass, herbaceous flowering plants present. e.g. milkweed, goldenrod, aster, etc. |

**Figure S1.**
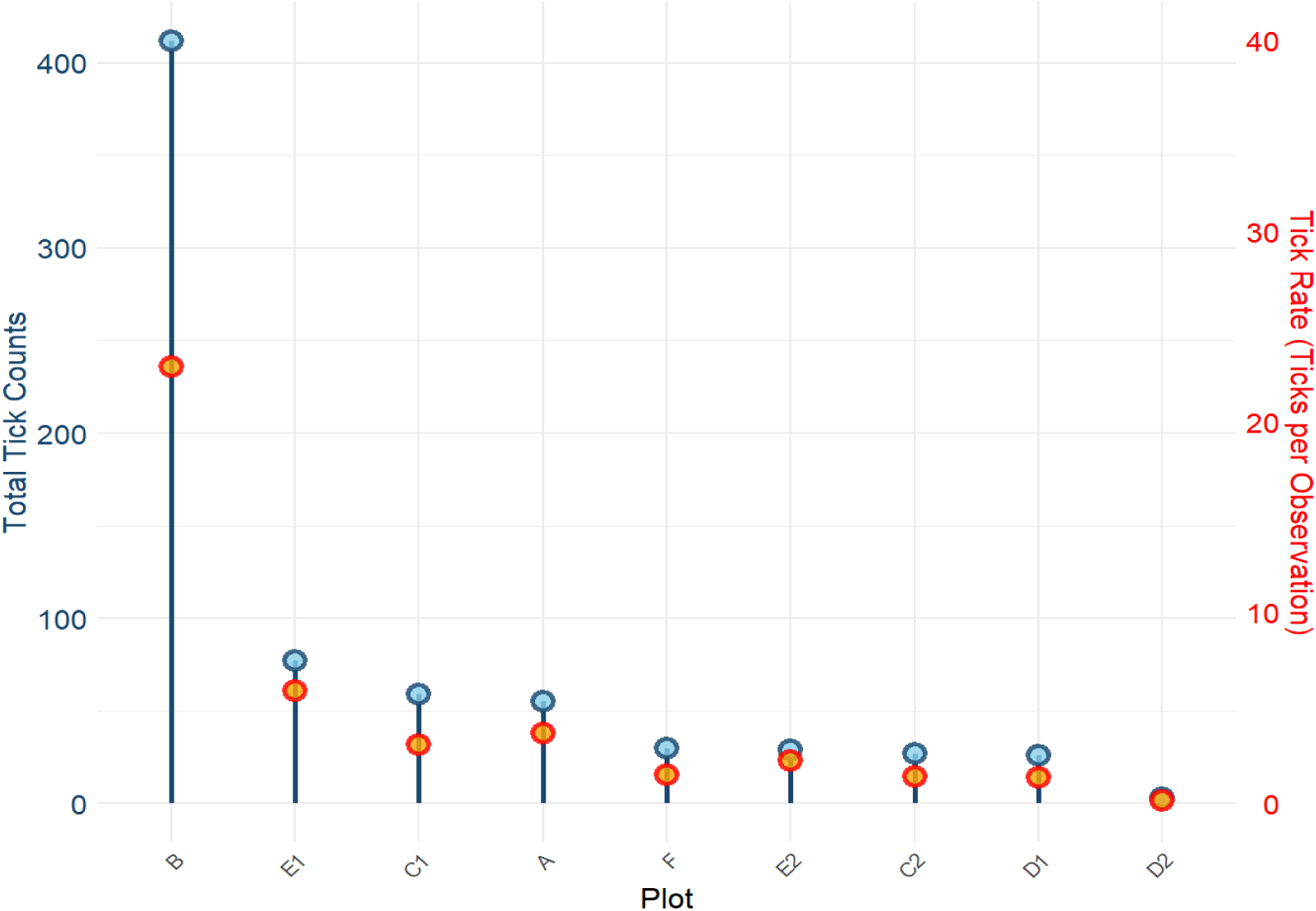
*Ixodes scapularis* totals and sampled densities, Western Massachusetts, 2024-2025.

**Table S4.** Summarized plot vegetation indices used for multivariable models for tick densities, Western Massachusetts, 2024-2025.

| Plot | Leaf Litter Depth (cm) | Leaf Litter Composition | Understory Vegetation Density (0 - 3) | Basal Area (m<sup>2</sup> / hectare) | Invasive Species Density | Percent Canopy Cover (%) | Mast Species | Berry Species | Coarse Woody Debris | Forbs | Short Brushy Veg. | Tall Brushy Veg. | Tall Grasses | Ferns | Height of Tallest Plant in Plot. |
| --- | --- | --- | --- | --- | --- | --- | --- | --- | --- | --- | --- | --- | --- | --- | --- |
| A | 4.53 | Mixed | 2.37 | 36.4 | 0 | 100 | 0.625 | 0.75 | 1.0 | 0.625 | 1.0 | 0.5 | 0 | 0.875 | >1m |
| B | 3.28 | Mixed | 2.0 | 18.9 | 1 | 62.5 | 0.625 | 1.0 | 0.875 | 1 | 1.0 | 0.5 | 0.75 | 0.125 | <0.5m |
| C1 | 3.15 | Deciduous | 2.37 | 30.4 | 0 | 100 | 1.0 | 1.0 | 0.5 | 0 | 1.0 | 1 | 0.25 | 0 | >1m |
| C2 | 2.67 | Deciduous | 1.62 | 43.0 | 0 | 100 | 1.0 | 0.375 | 0.625 | 0.25 | 1.0 | 0.75 | 0.5 | 0 | >1m |
| D1 | 3.65 | Deciduous | 1.87 | 25.5 | 0 | 100 | 1.0 | 0.375 | 0.625 | 0.25 | 1.0 | 1 | 0 | 0 | >1m |
| D2 | 2.83 | Mixed | 0.65 | 25.8 | 0 | 100 | 0.75 | 0 | 0.5 | 0.375 | 0.5 | 0.375 | 0.125 | 0 | >1m |
| E1 | 3.78 | Mixed | 2.37 | 28.7 | 0 | 100 | 0.875 | 0.5 | 0.75 | 0.625 | 0.625 | 0.625 | 0 | 0.5 | >1m |
| E2 | 3.58 | Coniferous | 2.12 | 32.7 | 0 | 100 | 0.5 | 0.25 | 0.625 | 0.5 | 0.75 | 0.75 | 0 | 1.0 | 0.5 - 1m |
| F | 1.54 | Deciduous | 2.12 | 29.5 | 0.5 | 100 | 0.625 | 0.375 | 0.75 | 0.875 | 0.875 | 0.25 | 0.5 | 0.5 | <0.5m |

**Figure S2.**
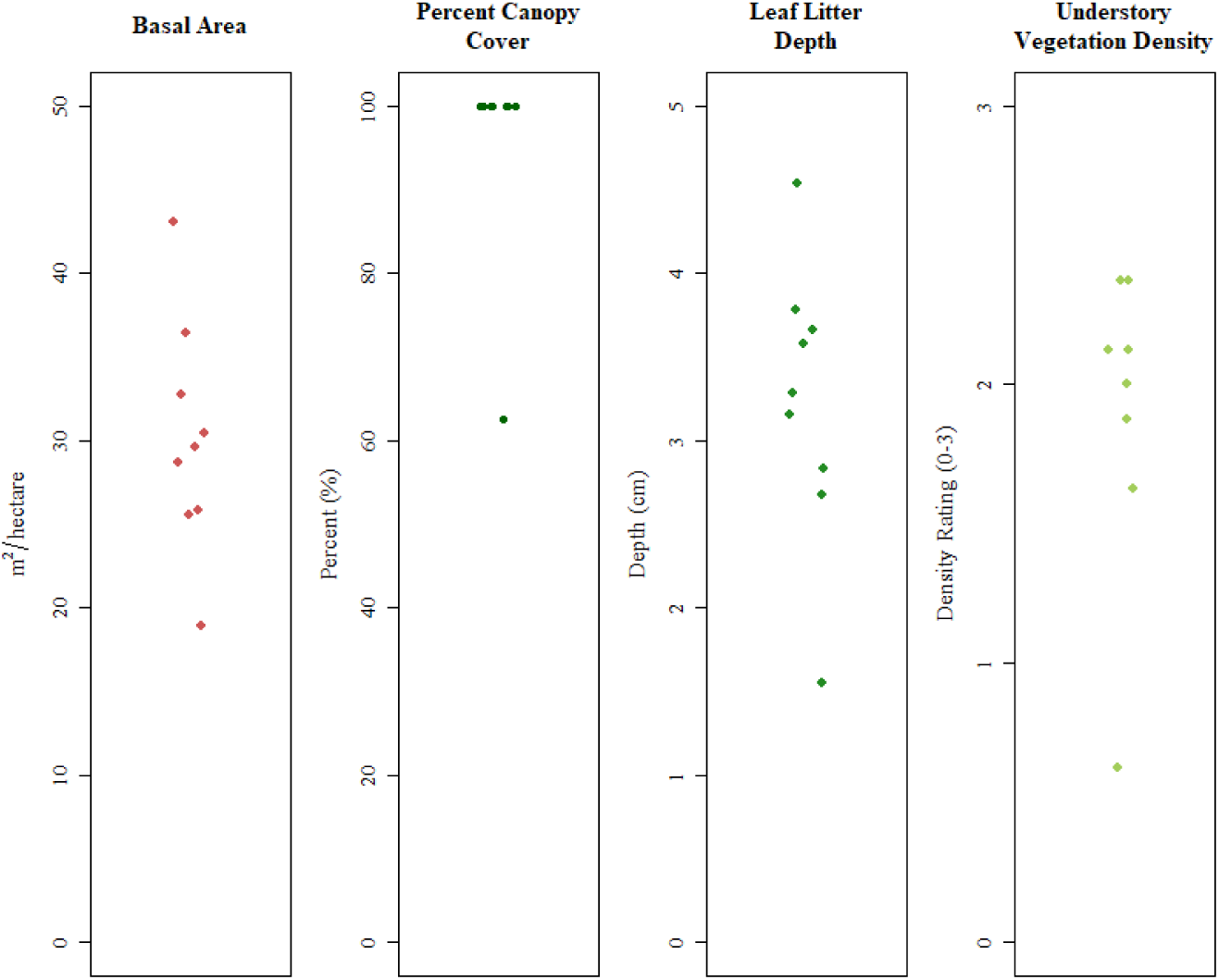
Overstory and understory composition strip plot of aggregated continuous and ordinal values observed from the vegetation survey, Western Massachusetts, 2024-2025.

**Figure S3.**
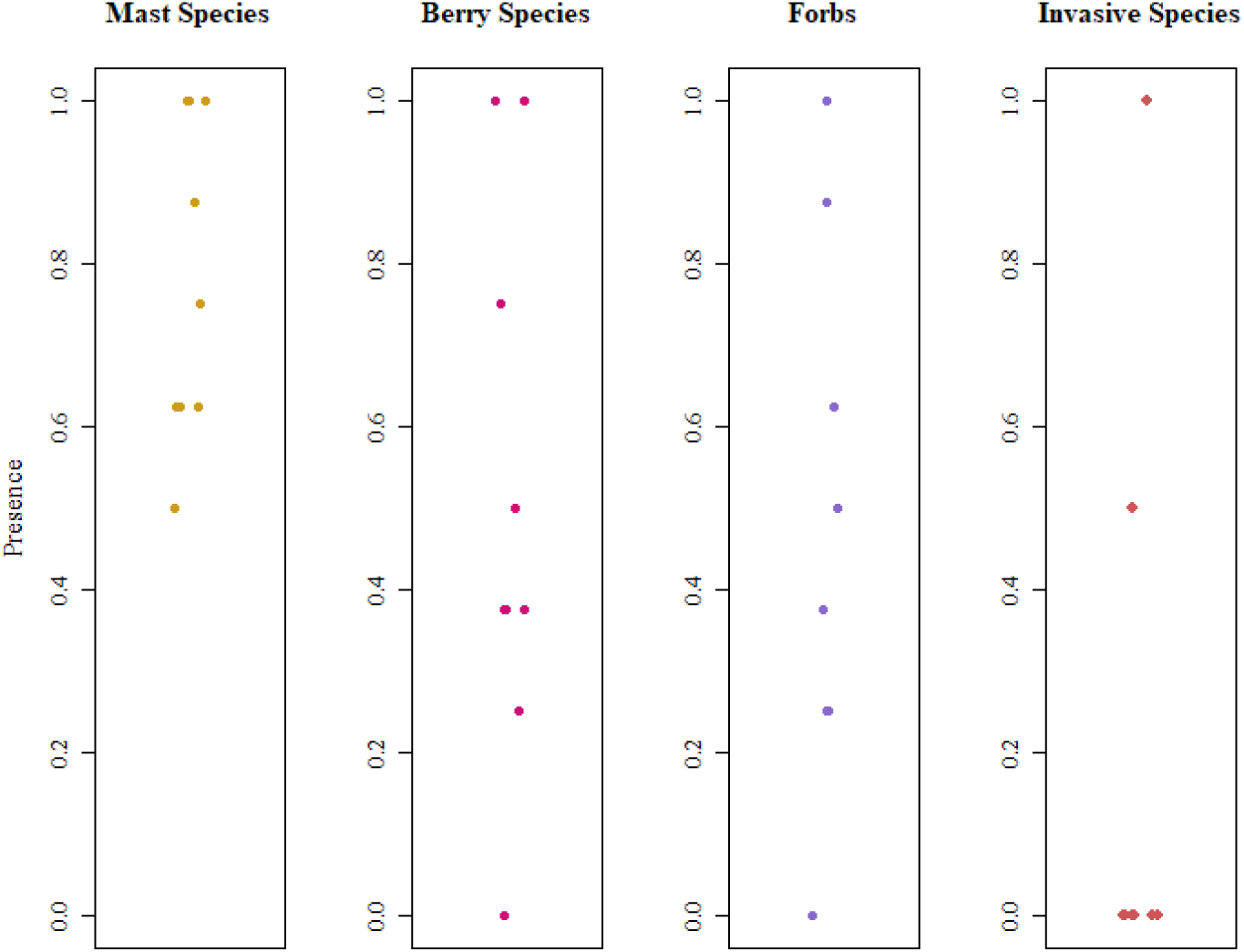
Plant species presence strip plot of averaged presence/absence values observed from the vegetation survey, Western Massachusetts, 2024-2025.

**Figure S4.**
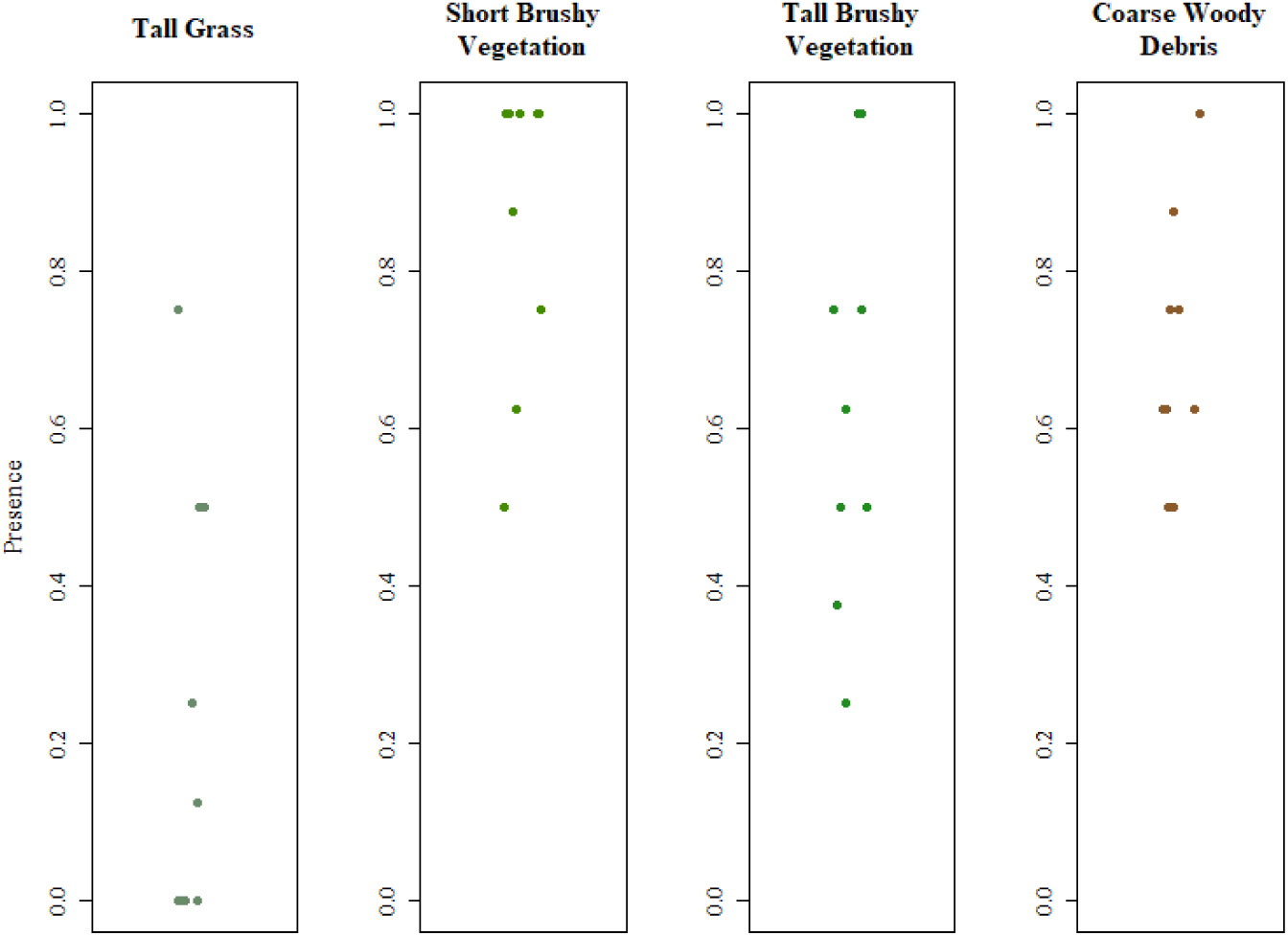
Understory composition strip plot of averaged presence/absence values observed from vegetation survey, Western Massachusetts, 2024-2025.

## Notes

### Author Declarations

IRB of University of Massachusetts Review Board gave ethical approval for this reviewed work. Approval #758 (Initial approval Mar 31, 2019)

## References

1 Rosenberg R, Lindsey NP, Fischer M, et al. *Vital Signs* : Trends in Reported Vectorborne Disease Cases — United States and Territories, 2004–2016. MMWR Morb Mortal Wkly Rep 2018; 67: 496–501.

2 Eisen L, Eisen RJ. Changes in the geographic distribution of the blacklegged tick, Ixodes scapularis, in the United States. Ticks Tick-Borne Dis 2023; 14: 102233.

3 Troughton DR, Levin ML. Life Cycles of Seven Ixodid Tick Species (Acari: Ixodidae) Under Standardized Laboratory Conditions. J Med Entomol 2007; 44: 732–40.

4 Sonenshine DE. Ecology of Non-nidiculous Ticks. In: Biology of Ticks Volume 2. New York: Oxford University Press, 1993: 14–8.

5 McBride SE, Lieberthal BA, Buttke DE, et al. Patterns and Ecological Mechanisms of Tick-Borne Disease Exposure Risk in Acadia National Park, Mount Desert Island, Maine, United States. J Med Entomol 2023; 60: 62–72.

6 Volk MR, Lubelczyk CB, Johnston JC, Levesque DL, Gardner AM. Microclimate conditions alter Ixodes scapularis (Acari: Ixodidae) overwinter survival across climate gradients in Maine, United States. Ticks Tick-Borne Dis 2022; 13: 101872.

7 Conte CE, Leahy JE, Gardner AM. Active Forest Management Reduces Blacklegged Tick and Tick-Borne Pathogen Exposure Risk. EcoHealth 2021; 18: 157–68.

8 Elias SP, Lubelczyk CB, Rand PW, Lacombe EH, Holman MS, Smith RP Jr. Deer Browse Resistant Exotic-Invasive Understory: An Indicator of Elevated Human Risk of Exposure to Ixodes scapularis (Acari: Ixodidae) in Southern Coastal Maine Woodlands. J Med Entomol 2006; 43: 1142–52.

9 Logan JJ, Knudby A, Leighton PA, et al. Ixodes scapularis density and Borrelia burgdorferi prevalence along a residential-woodland gradient in a region of emerging Lyme disease risk. Sci Rep 2024; 14: 13107.

10 Jordan RA, Schulze TL. Deer Browsing and the Distribution of *Ixodes scapularis* (Acari: Ixodidae) in Central New Jersey Forests. Environ Entomol 2005; 34: 801–6.

11 Lubelczyk CB, Elias SP, Rand PW, Holman MS, Lacombe EH, Smith RP Jr. Habitat Associations of Ixodes scapularis (Acari: Ixodidae) in Maine. Environ Entomol 2004; 33: 900–6.

12 Hurd SN, Kenefic LS, Leahy JE, Sponarski CC, Gardner AM. Cascading impacts of overstory structure in managed forests on understory structure, microclimate conditions, and *Ixodes scapularis* (Acari: Ixodidae) densities. J Med Entomol 2024; 61: 686–700.

13 Mathisson DC, Kross SM, Palmer MI, Diuk-Wasser MA. Effect of Vegetation on the Abundance of Tick Vectors in the Northeastern United States: A Review of the Literature. J Med Entomol 2021; 58: 2030–7.

14 Hook SA, Nelson CA, Mead PS. U.S. public’s experience with ticks and tick-borne diseases: Results from national HealthStyles surveys. Ticks Tick-Borne Dis 2015; 6: 483–8.

15 Tufa F, Ravenhurst J, Ballman ES, et al. Project ITCH: cross-sectional surveys to understand residential vector control attitudes and practices across New England. BMJ Public Health 2026; 4. DOI:10.1136/bmjph-2025-003656.

16 Shabani T, Jerie S, Shabani T. A comprehensive review of the Swiss cheese model in risk management. Saf Extreme Environ 2024; 6: 43–57.

17 Clark E, McFarland J, Parker B, Franklin S. The interface between invasive species and the increased incidence of tick-borne diseases, and the implications for federal land managers. National Invasive Species Council, 2019 https://digitalcommons.unl.edu/natlinvasive/33.

18 Catanzaro P, D’Amato A. Restoring Old-Growth Characteristics to New England’s and New York’s Forests. 2022. DOI:10.13140/RG.2.2.22226.84163.

19 Bellemare J, Motzkin G, Foster DR. Legacies of the agricultural past in the forested present: an assessment of historical land-use effects on rich mesic forests. J Biogeogr 2002; 29: 1401–20.

20 Butler BJ, Caputo J, Crocker S, et al. Southern New England Forests 2017: Summary Report. Resour Bull NRS-125 Madison WI US Dep Agric For Serv North Res Stn 15 P 2022; 125. DOI:10.2737/NRS-RB-125.

21 DeGraaf RM, Yamasaki M. New England Wildlife: Habitat, Natural History, and Distribution. Hanover, NH: University Press of New England, 2001.

22 Schulze TL, Jordan RA, Schulze CJ. Host Associations of Ixodes scapularis (Acari: Ixodidae) in Residential and Natural Settings in a Lyme Disease-Endemic Area in New Jersey. J Med Entomol 2005; 42: 966–73.

23 Halsey SJ, Allan BF, Miller JR. The role of *Ixodes scapularis*, Borrelia burgdorferi and wildlife hosts in Lyme disease prevalence: A quantitative review. Ticks Tick-Borne Dis 2018; 9: 1103–14.

24 Centers for Disease Control and Prevention. CDC Wonder. http://wonder.cdc.gov/.

25 Massachusetts Department of Public Health, Bureau of Infectious Disease and Laboratory Sciences. Tick Exposure and Tick-borne Disease Syndromic Surveillance Report, December 2022. http://www.mass.gov/eohhs/gov/departments/dph/programs/id/.

26 Massachusetts Census Data. https://malegislature.gov/Redistricting/MassachusettsCensusData/County (accessed Aug 14, 2026).

27 Tsalah J, Ravenhurst J, Snyder T, Bobskill V, Mahfuj A, Lover AA. Spatial Case Control Studies: Applying a Novel Study Design to Identify Potential Hotspots for Human Exposures to Ticks (Ixodidae) [Unpublished manuscript]. Department of Biostatistics and Epidemiology, University of Massachusetts Amherst. 2026.

28 Kulldorff M, Information Management Services, Inc. SaTScan. 2011. www.satscan.org.

29 Kleinman K. Rsatscan: tools, classes, and methods for interfacing with SaTScan stand-alone software. 2015; published online March 17. https://CRAN.R-project.org/package=rsatscan (accessed Feb 5, 2019).

30 Springer Y. TOS Protocol and Procedure: Tick and Tick-Borne Pathogen Sampling. NEON.DOC.014045VF. NEON (National Ecological Observatory Network). 2015; published online March 17.

31 Durden LA, Keirans JE. Key to the Nymphs of the Genus Ixodes of the United States. In: Nymphs of the Genus Ixodes (Acari: Ixodidae) of the United States: Taxonomy, Identification Key, Distribution, Hosts, and Medical/Veterinary Importance. Entomological Society of America, 1996: 5.

32 Keirans JE, Litwak TR. Pictorial Key to the Adults of Hard Ticks, Family Ixodidae (Ixodida: Ixodoidea), East of the Mississippi River. J Med Entomol 1989; 26: 435–48.

33 Light Grey Canvas. 2015; published online Nov 13. https://basemaps.arcgis.com/arcgis/rest/services/World_Basemap_v2/VectorTileServer.

34 Hahsler M, Buchta C, Gruen B, et al. arules: Mining Association Rules and Frequent Itemsets. 2026; published online March 29. https://cran.r-project.org/web/packages/arules/index.html (accessed June 30, 2026).

35 Brooks ME, Kristensen K, Benthem KJ van, et al. glmmTMB Balances Speed and Flexibility Among Packages for Zero-inflated Generalized Linear Mixed Modeling. R J 2017; 9: 378–400.

36 Brooks M, Bolker B, Kristensen K, et al. glmmTMB: Generalized Linear Mixed Models using Template Model Builder. 2025; published online Oct 9. https://cran.r-project.org/web/packages/glmmTMB/index.html (accessed Oct 21, 2025).

37 Hosmer DW, Lemeshow S. Applied logistic regression, 2nd edn. Wiley-Interscience Publication, 2000.

38 Lüdecke D, Ben-Shachar M, Patil I, Waggoner P, Makowski D. performance: An R Package for Assessment, Comparison and Testing of Statistical Models. J Open Source Softw 2021; 6: 3139.

39 Johnston R, Jones K, Manley D. Confounding and collinearity in regression analysis: a cautionary tale and an alternative procedure, illustrated by studies of British voting behaviour. Qual Quant 2018; 52: 1957–76.

40 Mansournia MA, Geroldinger A, Greenland S, Heinze G. Separation in Logistic Regression: Causes, Consequences, and Control. Am J Epidemiol 2018; 187: 864– 70.

41 Hartig F, Lohse L, Leite M de S, Werneke C. DHARMa: Residual Diagnostics for Hierarchical (Multi-Level / Mixed) Regression Models. 2026; published online June 1. https://cran.r-project.org/web/packages/DHARMa/index.html (accessed July 17, 2026).

42 Yoder JA, Spielman A. Differential capacity of larval deer ticks (*Ixodes dammini*) to imbibe water from subsaturated air. J Insect Physiol 1992; 38: 863–9.

43 Brunner JL, LaDeau SL, Killilea M, Valentine E, Schierer M, Ostfeld RS. Off-host survival of blacklegged ticks in eastern North America: A multistage, multiyear, multisite study. Ecol Monogr 2023; 93: e1572.

44 Schulze TL, Jordan RA. Influence of meso-and microscale habitat structure on focal distribution of sympatric Ixodes scapularis and Amblyomma americanum (Acari: Ixodidae). J Med Entomol 2005; 42: 285–94.

45 Carroll JF, Schmidtmann ET. Dispersal of Blacklegged Tick (Acari: Ixodidae) Nymphs and Adults at the Woods–Pasture Interface. J Med Entomol 1996; 33: 554– 8.

46 Schulze TL, Jordan RA. Influence of Meso- and Microscale Habitat Structure on Focal Distribution of Sympatric Ixodes scapularis and Amblyomma americanum (Acari: Ixodidae). J Med Entomol 2005; 42: 285–94.

47 Brennan RN, Boychuck S, Washkwich AJ, John-Alder H, Fonseca DM. Tick abundance and diversity are substantially lower in thinned vs. unthinned forests in the New Jersey Pinelands National Reserve, USA. Ticks Tick-Borne Dis 2023; 14: 102106.

48 Bespyatova LА, Bugmyrin SV. Ixodid ticks (Parasitiformes: Ixodidae) from small mammals in reforesting boreal habitats of the northern European Russia. Entomol Rev 2015; 95: 1308–16.

49 Ehrmann S, Ruyts SC, Scherer-Lorenzen M, et al. Habitat properties are key drivers of Borrelia burgdorferi (s.l.) prevalence in Ixodes ricinus populations of deciduous forest fragments. Parasit Vectors 2018; 11: 23.

50 Williams SC, Linske MA, Ward JS. Long-Term Effects of Berberis thunbergii (Ranunculales: Berberidaceae) Management on Ixodes scapularis (Acari: Ixodidae) Abundance and Borrelia burgdorferi (Spirochaetales: Spirochaetaceae) Prevalence in Connecticut, USA. Environ Entomol 2017; 46: 1329–38.

51 Lindström A, Jaenson TGT. Distribution of the Common Tick, *Ixodes ricinus* (Acari: Ixodidae), in Different Vegetation Types in Southern Sweden. J Med Entomol 2003; 40: 375–8.

52 Kollars TM Jr, Oliver JH Jr, Kollars PG, Durden LA. Seasonal Activity and Host Associations of Ixodes scapularis (Acari: Ixodidae) in Southeastern Missouri. J Med Entomol 1999; 36: 720–6.

53 Baldwin H, Landesman WJ, Borgmann-Winter B, Allen D. A Geographic Information System Approach to Map Tick Exposure Risk at a Scale for Public Health Intervention. J Med Entomol 2022; 59: 162–72.

54 Lambin EF, Tran A, Vanwambeke SO, Linard C, Soti V. Pathogenic landscapes: Interactions between land, people, disease vectors, and their animal hosts. Int J Health Geogr 2010; 9: 54.

55 Ginsberg HS, Rulison EL, Miller JL, et al. Local abundance of *Ixodes scapularis* in forests: Effects of environmental moisture, vegetation characteristics, and host abundance. Ticks Tick-Borne Dis 2020; 11: 101271.

56 Hurd SN, Kenefic LS, Leahy JE, Sponarski CC, Gardner AM. Cascading impacts of overstory structure in managed forests on understory structure, microclimate conditions, and Ixodes scapularis (Acari: Ixodidae) densities. J Med Entomol 2024; 61: 686–700.

57 Diuk-Wasser MA, Vourc’h G, Cislo P, et al. Field and climate-based model for predicting the density of host-seeking nymphal Ixodes scapularis, an important vector of tick-borne disease agents in the eastern United States. Glob Ecol Biogeogr 2010; 19: 504–14.

58 Tick Knowledge, Surveillance, Control, and Communication Efforts among Midwestern and Northeastern Land Trusts. https://scholarworks.iu.edu/iuswrrest/api/core/bitstreams/ec999915-ddf5-40f8-83ee-94dd65675f81/content.

59 Audet-Legault R, Hainault J, Desrochers-Plourde C-A, et al. Effectiveness of tick sampling methods in the field: A systematic scoping review. Ticks Tick-Borne Dis 2026; 17: 102682.

60 Mandli JT, Lee X, Bron GM, Paskewitz SM. Integrated Tick Management in South Central Wisconsin: Impact of Invasive Vegetation Removal and Host-Targeted Acaricides on the Density of Questing *Ixodes scapularis* (Acari: Ixodidae) Nymphs. J Med Entomol 2021; 58: 2358–67.

61 Elmieh N. Review of environmental management strategies to reduce tick populations. Vancouver, BC: Canadian National Collaborating Centre for Environmental Health, 2023 https://ncceh.ca/resources/evidence-reviews/review-environmental-management-strategies-reduce-tick-populations.

62 Quine CP, Barnett J, Dobson ADM, et al. Frameworks for risk communication and disease management: the case of Lyme disease and countryside users. Philos Trans R Soc B Biol Sci 2011; 366: 2010–22.

63 Brady RM, Lemieux CJ, Doherty ST. Ticks and Lyme disease in natural areas: A segmentation analysis of visitor perceptions of risk and preferred communication strategies. J Outdoor Recreat Tour 2024; 47: 100794.

64 Dumas A, Bouchard C, Drapeau P, Lindsay LR, Ogden NH, Leighton PA. The risk of contact between visitors and Borrelia burgdorferi-infected ticks is associated with fine-scale landscape features in a southeastern Canadian nature park. BMC Public Health 2024; 24: 1180.

65 Rochlin I, Ninivaggi DV, Benach JL. Malaria and Lyme disease - the largest vector-borne US epidemics in the last 100 years: success and failure of public health. BMC Public Health 2019; 19: 804.

