## Supplementary material for "Quantifying Fine-scale Vegetation Associated with *Ixodes scapularis* Density to Inform Public Health Programming, Western Massachusetts": Vegetation Survey Collection Sheet

### SEHTI 2024 Vegetation Survey

*Sampling a 3x3m subplot every 20m*

**Site ID** ______________________________ **Plot** ___________________________ **Subplot #**______

**Date assessed** ____________________ **Assessed by**____________

Leaf Litter present? (circle): **Yes** or **No**

| | |
|---|---|
| Depth: cm | Deciduous / Coniferous / Mixed (circle) |
| Depth: cm | |

| **Invasive species density: 0 1 2 3** | |
|---|---|
| *If invasive species density scored 1 or above, select species seen* | |
| ❑ Asiatic bittersweet | ❑ Multiflora rose |
| ❑ Barberry (Japanese and common) | ❑ Norway maple |
| ❑ Black locust | ❑ Purple loosestrife |
| ❑ Bush Honeysuckle | ❑ Winged Euonymus |
| ❑ Garlic mustard | ❑ ____________________________ |
| ❑ Glossy Buckthorn | ❑ ____________________________ |
| ❑ Common Buckthorn | ❑ ____________________________ |

| | |
|---|---|
| Forest floor vegetation coverage | **0 1 2 3** |
| Height of tallest plant/shrub in plot | **None <0.5m 0.5-1m >1m** |
| Basal Area (BAF:10) | Tally:<br>_____(total number) x 10 = [ ] sq ft/ac |
| Canopy Cover<br>*Formula* | *Mark presence of canopy at each point* |

Canopy Cover formula:

| Hits | 1 | 2 | 3 | 4 | 5 | 6 | 7 | 8 |
|---|---|---|---|---|---|---|---|---|
| %CC | 13 | 25 | 38 | 50 | 63 | 75 | 88 | 100 |

Mark presence of canopy at each point:

| 1 | 2 | 3 | 4 | 5 | 6 | 7 | 8 | Σ |
|---|---|---|---|---|---|---|---|---|

Canopy Cover= [ % ]

| **Mark presence of the following categories** | |
|---|---|
| ❑ Mast Species (Oak, Hickory, Walnut) | ❑ **TALL** brushy vegetation |
| ❑ Berry species (Vaccinium, Rubus) | ❑ Tall grasses |
| ❑ Coarse woody debris | ❑ Forbs |
| ❑ **short** brushy vegetation | ❑ Ferns |

NOTES:

______________________________________________________________________________

### SEHTI 2024 Vegetation Survey

*Sampling a 3x3m subplot every 20m*

KEY

*Mast Species*- Oak, hickory or walnut species of any age within the subplot (either leaning in and providing canopy coverage, or growing within the subplot)

*Berry species*- vaccinium and Rubus species present within or leaning into the subplot

*Coarse woody debris*– downed trees or large branches (at least 10cm in diameter and 1m in length)

*Short brushy vegetation* - dense cover of short (<2.5ft) shrubs that are easily passable

*Tall brushy vegetation* - dense cover of tall shrubs which obstructs free passage. May require flagging

*Tall grasses* - grasses, rushes, sedges (graminoids) and other weeds measuring 8 inches or greater in height

*Ground cover vegetation* - low-growing vegetation <3ft, fully covers ground in **50% or more** of area being evaluated

*Forbs* – non-grass, herbaceous flowering plants present. E.g. Milkweed, Goldenrod, Aster, etc.
