## Supplementary material for "Quantifying Fine-scale Vegetation Associated with *Ixodes scapularis* Density to Inform Public Health Programming, Western Massachusetts": Vegetation Survey Protocol

### Long-Term Tick Surveillance Sites (WMA) – Vegetation Survey Protocol

#### Summary

This guide provides an overview for conducting vegetation surveys in 3x3 subplots spaced 20 meters apart. It outlines methods for measuring leaf litter, assessing vegetation and invasive species density, recording basal area, and categorizing plant height. The guide also covers canopy coverage assessment, as well as the presence or absence of key species like mast trees, berry species, and other notable features.

Please contact Johanna Ravenhurst and Veronica Bobskill with any questions.

### I. Overview

#### Goals

Quantify the inter-annual variability in local tick populations, at a wide range of different ecological sites.

- Establish long-term tick surveillance sites in the Pioneer Valley
- Use spatial epidemiology to select potential sites based on human activity hotspot analysis
- Use NEON protocols to conduct active tick surveillance every 2 weeks
- Use hobo dataloggers to track temperature and humidity at sites.
- Design & conduct vegetation analyses at all sites, adapting protocols from NEWVEC collaborators in VT and ME. Aim to conduct analyses that have results that can be compared with collaborator data.

#### Site Selection

Potential sites were identified using a hotspot analysis from an NIH-funded spatial case-control study. Hotspots near trailheads were considered for long-term surveillance. Ideally, we are looking for sites with frequent human use. Beyond the hotspots, we are considering a few other potential locations around Western MA.

Exclusion criteria include:

- lack of sufficient tick habitat to set up a plot
- wetland
- widespread poison ivy
- thick thorny plants that can't be avoided for sampling.

As of September 2024, we have 4 sites and a total of 6 plots finalized. We are evaluating additional sites, and plan to add up to 6 more sites for long-term surveillance.

#### Tick Sampling

We are sampling a 160m perimeter (typically a 40x40m square). Sampling is conducted every 2 weeks, using a standard 1m$^2$ rubberized flannel drag cloth and stopping every 10 meters to collect ticks. Flagging is conducted with the same cloth in taller vegetation, such as mountain laurel. Ticks are removed from the site for identification and pathogen testing.

#### Vegetation Sampling

Vegetation sampling is conducted at our tick surveillance plots. Every 20 meters, a 3x3m subplot is created along the edge of the tick sampling plot, for evaluating the vegetation. There will be a total of 8 subplots per tick surveillance plot, or up to 16 per site.

Vegetation Data Collection Summary:

1. Leaf litter depth and composition
    a. Coniferous vs Deciduous
2. Invasive species density (0-3 scale)
    a. Invasive species will be noted
3. Understory vegetation coverage (0-3)
    a. Includes both invasive and noninvasive species present
4. Plant height (<0.5m, 0.5-1m, >1m)
    a. Includes plants growing within the subplot and/or leaning into the subplot. It would only include the plants a deer could feasibly lean into and get a tick from.
5. Basal area
    a. Basal area would be collected from the center of the subplot using a BAF of 10
6. Canopy coverage using a GRS Densitometer
7. Presence/Absence
    a. Mast Species (Oak, Hickory, Walnut)
    b. Berry species (Vaccinium, Rubus)
    c. Coarse woody debris
    d. short brushy vegetation (<2.5ft)
    e. TALL brushy vegetation (>2.5ft)
    f. Tall grasses
    g. Forbs
    h. Ferns

#### Comparing with Collaborator Methods

Each state has different tick collecting methods and variation in vegetation. This protocol is based largely on methods used by Vermont collaborators, Dr. Kristen Ross and Dr. William Landesman. Their tick surveillance is slightly different from our plots because it occurs at random points and consists of belt transects. Our field staff travelled to Vermont to learn about their vegetation survey methods.

Ross and Landesman use 3 x 3 meter subplots for vegetation surveys. These subplots are placed at randomly generated points around the tick surveillance belt transect. We are also using 3 x 3 meter subplots, but the plots are located along the edge of the plot. This makes it faster to identify the subplot locations, and we are analyzing the vegetation along the transects where we sample for ticks. However, we are not analyzing subplots within the tick surveillance plot, which could limit our ability to generalize vegetation analysis findings to the entire site.

We kept some of the same measurements as Ross and Landesman's vegetation analysis, such as the invasive species density and the height of tallest plant/shrub in plot.

We also learned from past vegetation surveys by our Maine collaborator, Dr. Allison Gardner. We consulted several publications (https://doi.org/10.1093/jme/tjac152 & https://doi.org/10.1093/jme/tjae030), and discussed our vegetation protocol with Dr. Gardner. We kept some of the same measurements as Gardner's vegetation protocols, such as percent canopy cover, and leaf little depth and composition.

We also discussed our protocol with UMass collaborator Dr. Allison Snow, who has conducted tick surveillance and vegetation surveys on Martha's Vineyard and Nantucket. We discussed the factors that might have the most impact on tick populations, such as shade, humidity, shrub layer, leaf litter depth, and proximity to wetland. This helped us to narrow down our list of potential vegetation measurements.

#### II. Sampling

##### Surveying Protocol Overview

Sampling will occur from August-September during larval deer tick season. Surveying will not occur in heavy rain but may in some mildly moist conditions.

Number of Sampling Locations (As of August 2024):

- 4 sites in the Pioneer Valley
    - Mt Tom
    - Leeds
    - Lowland Loop

  - K.C. Trail
- 1-2 tick surveillance plots per site

#### Data Collection

##### Step 1 – Creating a subplot

*Materials needed: Flagging tape*

*A.* Starting at the SW corner of the plot, measure 18.5 meters clockwise, following the edge.
B. Using the 3m PVC pipe, outline a 3x3m subplot, placing flags/chaining pins on each corner. The center of this plot should lie 20m from the starting point.

Each subsequent plot center should be 20 meters from the previous plot center.

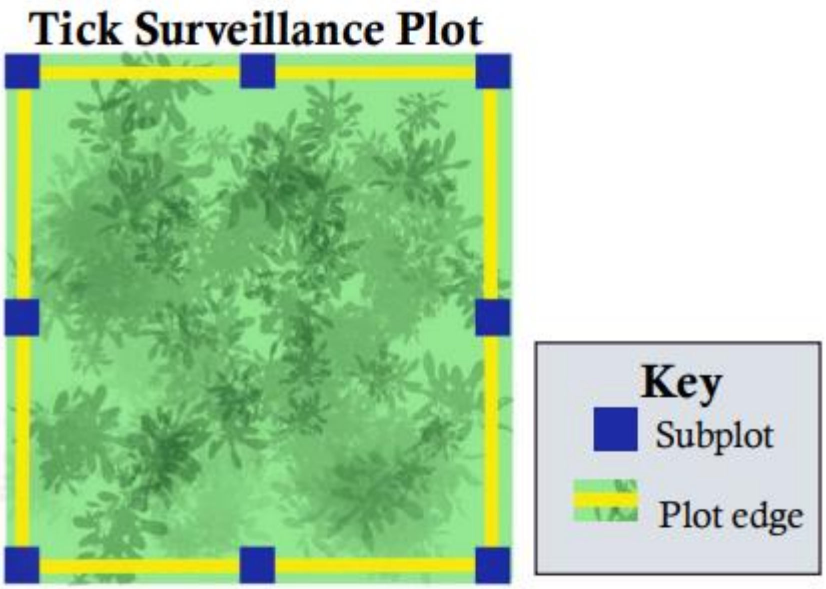

##### Step 2 – Vegetation Survey Sheet Header

*Materials needed: Vegetation Collection Data Sheet (Appendix C), writing implements, storage clipboard*

A. Fill in header section of the *Vegetation Collection Data Sheet*
   a. Subplot number follows the order of sampling for each plot, beginning with 1 and ending at 8.

SEHTI 2024 Vegetation Survey
*Sampling a 3x3m subplot every 20m*

**Site ID** ________________________ **Plot** _____________________ **Subplot #**_____

**Date assessed** ________________ **Assessed by**__________

#### Step 3 – Vegetation Survey

##### 1. Leaf litter depth and composition

*Materials needed: Hori-Hori Soil knife, ruler*

**If marked present,** leaf litter composition should be assessed as one of the three categories on the datasheet: Deciduous, Coniferous, and Mixed

*Deciduous: >75% of material from deciduous vegetation*

*Coniferous: >75% of material from coniferous vegetation*

*Mixed: roughly even mix of material from both deciduous and coniferous types*

Survey the left and right side of your subplot in the center of each edge.

Using your hori-hori knife, plunge into the soil and wiggle front to back until there is a small gap where a ruler can be used. Measure in centimeters and repeat on the other side.

##### 2. Invasive species density

Invasive species density is described on a 0-3 scale, with 0 being none present, and 3 being entirely filled. There will likely be non-invasive species present concurrently, ignore all native plants when scaling invasive presence.

| Invasive Density: 1 | Invasive Density: 2 | Invasive Density: 3 |
| --- | --- | --- |

| | | |
|---|---|---|
| 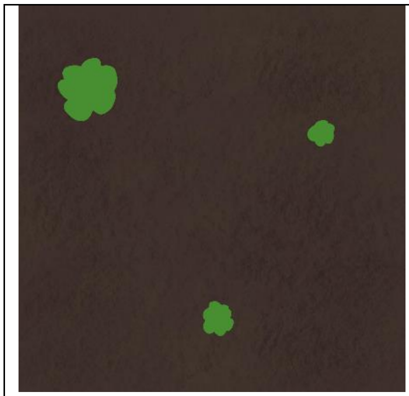 | 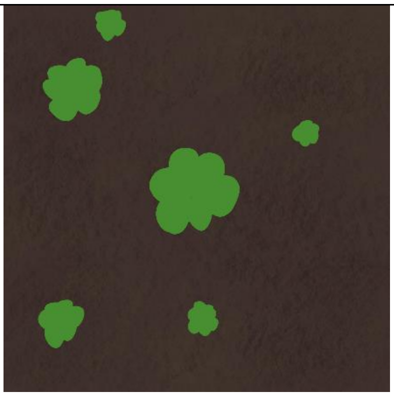 | 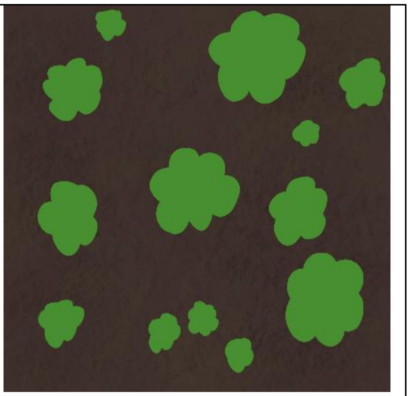 |

A value of 3 could be a few large individuals that fill up a subplot, or many small.

#### 3. Understory vegetation coverage

Understory vegetation coverage includes both native and non-native species that make up the lower forest. This is described on a 0-3 scale, with 0 being none present, and 3 being entirely filled.

| Vegetation coverage: 1 | Vegetation coverage: 2 | Vegetation coverage: 3 |
|---|---|---|
| 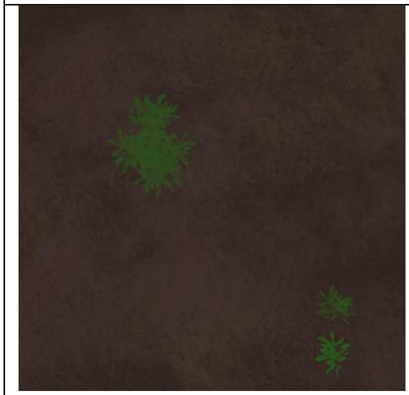 | 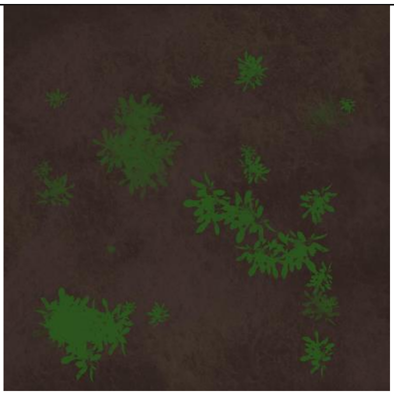 | 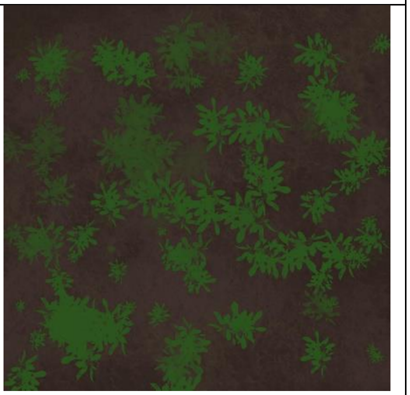 |

#### 4. Plant Height

*Materials needed: 60m/200ft Measuring Tape*

Plant height in this assessment refers only to the tallest plant within the plot, or leaning into the plot that mammals, such as deer, could feasibly brush against. Use the measuring tape to determine what height range this plant falls into: <0.5m, 0.5-1m, >1m

#### 5. Basal area

*Materials needed: Cruz-All*

Stand in the center of the plot. Hold the Cruz-All at eye height, making sure the attached chain is tight when you do so. Slowly complete a 360° turn, counting the number of "in" trees with a BAF (Basal Area Factor) 10. Any tree spanning the entire opening of the Cruz-All at BAF 10 is counted as in.

Complete the Basal Area section of the Vegetation Survey sheet by inputting the total count of "in" trees and multiplying by 10 to find basal area.

#### 6. Canopy coverage

*Materials needed: GRS Densitometer*

To use a GRS densitometer, it must be held a small distance (<6 inches) from your face. There are two levels inside, and each must have a bubble in the center before your measurement is collected.

After the densitometer is determined to be even, move your head until the center circle is in the center of the crosshairs. Once you are properly placed, you may determine if there is vegetation in your circle. Even if there is a small amount of canopy in your circle, it will be marked as "yes"

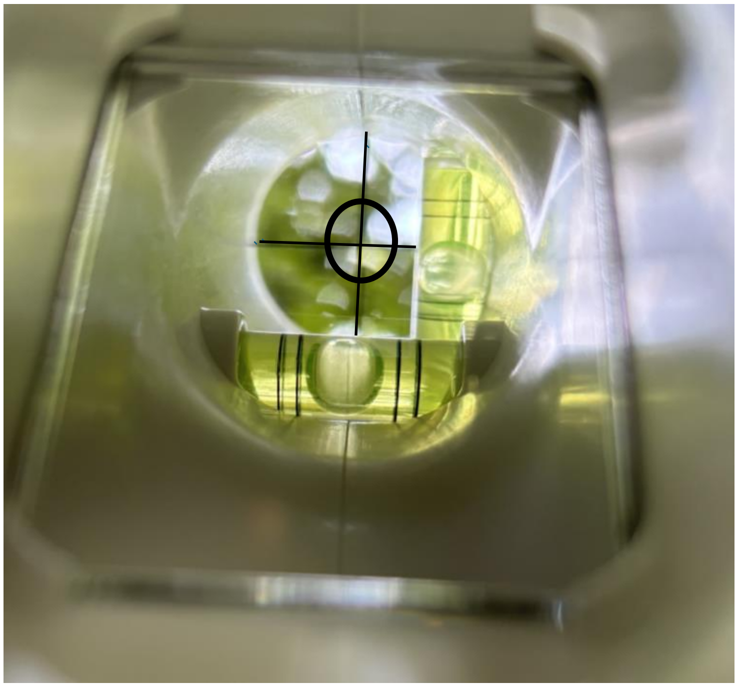

See photo on the right for the view within a densitometer. *Note: Vegetation is clearer during actual use, but cameras*

*struggle to focus both crosshairs and vegetation.*

Using the following formula, canopy coverage can be calculated.

$$Percent\ canopy\ cover = \left(\frac{\#\ \text{yes}\ hits}{total\ observations}\right) x\ 100$$

To determine canopy cover on the subplot, 8 points will be taken. One on each corner, and one between each corner. Based on the total of “yes” canopy hits, percent canopy cover can be calculated. Using the following key, with values rounded to whole numbers, there is no in-field math necessary.

| “yes” hits | 1 | 2 | 3 | 4 | 5 | 6 | 7 | 8 |
| --- | --- | --- | --- | --- | --- | --- | --- | --- |
| % canopy cover | 13 | 25 | 38 | 50 | 63 | 75 | 88 | 100 |

##### 7. Presence/Absence

Note the presence of the following features present within the subplot

- Mast Species (Oak, Hickory, Walnut)
- Berry species (*Vaccinium, Rubus*)
- Coarse woody debris
- short brushy vegetation
- TALL brushy vegetation
- Tall grasses
- Forbs
- Ferns

See **Appendix A** for further explanation on presence/absence and **Appendix B** for a full materials list.

#### III. Data Management & Quality Control

##### Paper Form Data Entry

Paper forms will be used for the vegetation survey and other key information while sampling in the field. All data captured on paper forms should be entered in Qualtrics within 2 weeks of collection. Ideally this should be completed by the person(s) who collected the data. A scan/picture of the paper form should also be uploaded to Qualtrics as a backup.

See **Appendix C** for data sheet used.

#### Appendix A: Presence/Absence Glossary

*Mast Species*- Oak, hickory or walnut species of any age within the subplot (either leaning in and providing canopy coverage or growing within the subplot). If you cannot determine whether a tree is leaning into the plot, use the GRS densitometer to look directly above.

*Berry species*- Vaccinium and Rubus species present within or leaning into the subplot

*coarse woody debris*– downed trees or large branches (at least 10cm in diameter and 1m in length)

*Short brushy vegetation* - dense cover of short (<2.5ft) shrubs that are easily passable

*Tall brushy vegetation* - dense cover of tall shrubs which obstructs free passage. May require flagging

*tall grasses* - grasses, rushes, sedges (graminoids) and other weeds measuring 8 inches or greater in height

*ground cover vegetation* - low-growing vegetation <3ft, fully covers ground in **50% or more** of area being evaluated

*forbs* – non-grass, herbaceous flowering plants present. E.g. Milkweed, Goldenrod, Aster, etc.

#### Appendix B: Materials

| **Supplier/<br>Item Number** | **Item** | **Purpose** | **Image** |
|---|---|---|---|
| Forestry Suppliers - Growtech | Hori-Hori Soil knife | To find leaf litter depth | 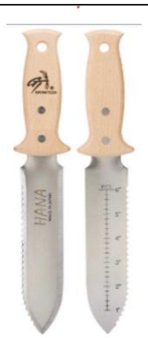 |
| Keson - Metric/SAE - 200 ft/60m<br>Grainger - Item # 3LJT5 | 60m/200ft Measuring Tape | Measures distance between subplots, to measure out the subplots, and to determine height of vegetation within the subplot | 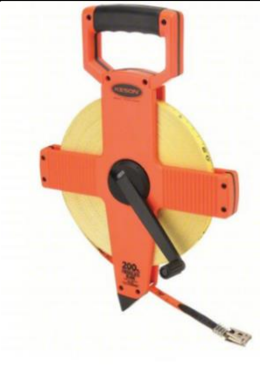 |
| Home Depot or Forestry Suppliers | Flagging tape (pink color for survey marks) OR chaining pins | Mark edges of plot | 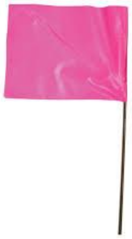<br>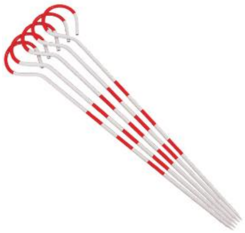 |

| | | | |
|---|---|---|---|
| Forestry Suppliers | GRS Densitometer | Survey canopy closure | 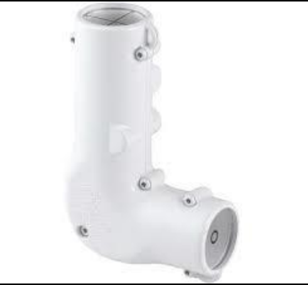 |
| N/A | Storage clipboard | Holds survey data sheets and writing implements | 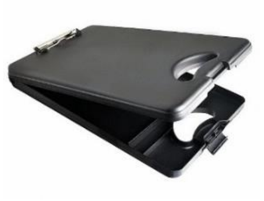 |
| Forestry suppliers | Jim-Gem Cruz-All | Find basal area | 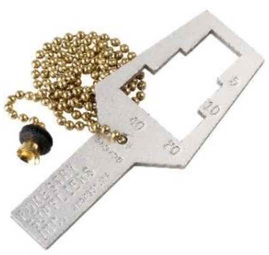 |
| | 3m PVC pipe | Quickly measure subplot | |
| W.B Mason | Ruler | Measure leaf litter depth | |
| Veronica Bobskill and Mark Titus | Vegetation survey data collection sheet | Record information | |

#### Appendix C: Vegetation Collection Data Sheet

WMA 2024 Vegetation Survey

*Sampling a 3x3m subplot every 20m*

**Site ID** ____________________________ **Plot** _________________________ **Subplot #**______

**Date assessed** ___________________ **Assessed by**____________

Leaf Litter present? (circle): **Yes** or **No**

| Depth: cm | Deciduous / Coniferous / Mixed (circle) |
|---|---|
| Depth: cm | |

| **Invasive species density: 0 1 2 3** | |
|---|---|
| *If invasive species density scored 1 or above, select species seen* | |
| ❑ Asiatic bittersweet | ❑ Multiflora rose |
| ❑ Barberry (Japanese and common) | ❑ Norway maple |
| ❑ Black locust | ❑ Purple loosestrife |
| ❑ Bush Honeysuckle | ❑ Winged Euonymus |
| ❑ Garlic mustard | ❑ ___________________________ |
| ❑ Glossy Buckthorn | ❑ ___________________________ |
| ❑ Common Buckthorn | ❑ ___________________________ |

| Forest floor vegetation coverage | **0 1 2 3** |
|---|---|
| Height of tallest plant/shrub in plot | **None <0.5m 0.5-1m >1m** |
| Basal Area (BAF:10) | Tally:<br>_____(total number) x 10 = ☐ sq ft/ac |

Canopy Cover

*Formula*

| Hit | 1 | 2 | 3 | 4 | 5 | 6 | 7 | 8 |
|---|---|---|---|---|---|---|---|---|
| %CC | 13 | 25 | 38 | 50 | 63 | 75 | 88 | 100 |

*Mark presence of canopy at each point*

| 1 | 2 | 3 | 4 | 5 | 6 | 7 | 8 | Σ |
|---|---|---|---|---|---|---|---|---|

Canopy Cover= ______ %

| Mark presence of the following categories | |
|---|---|
| ❑ Mast Species (Oak, Hickory, Walnut) | ❑ **TALL** brushy vegetation |
| ❑ Berry species (Vaccinium, Rubus) | ❑ Tall grasses |
| ❑ Coarse woody debris | ❑ Forbs |
| ❑ **short** brushy vegetation | ❑ Ferns |

NOTES:

________________________________________

________________________________________

________________________________________

________________________________________
